# Networks of placental DNA methylation correlate with maternal serum PCB concentrations and child neurodevelopment

**DOI:** 10.1101/2022.11.13.22282272

**Authors:** Julia S. Mouat, Xueshu Li, Kari Neier, Yihui Zhu, Charles E. Mordaunt, Michele A. La Merrill, Hans-Joachim Lehmler, Michael P. Jones, Pamela J. Lein, Rebecca J. Schmidt, Janine M. LaSalle

## Abstract

**Background:** Gestational exposure to polychlorinated biphenyls (PCBs) has been associated with elevated risk for neurodevelopmental disorders. The mechanism of risk is unclear but may involve placental epigenetics. Prior studies have associated differential placental DNA methylation with maternal PCB exposure or with increased risk of autism spectrum disorder (ASD). However, sequencing-based placental methylomes have not previously been tested for simultaneous associations with maternal PCB levels and child neurodevelopmental outcomes.

**Objectives:** We aimed to identify placental DNA methylation patterns associated with maternal PCB levels and child neurodevelopmental outcomes in the high-risk ASD MARBLES cohort.

**Methods:** We measured 209 PCB congeners in 104 maternal serum samples collected at delivery. We identified networks of DNA methylation from 147 placenta samples using the Comethyl R package, which performs weighted gene correlation network analysis for whole genome bisulfite sequencing data. We tested placental DNA methylation modules for association with maternal serum PCB levels, child neurodevelopment, and other participant traits.

**Results:** PCBs 153 + 168, 170, 180 + 193, and 187 were detected in over 50% of maternal serum samples and were highly correlated with one another. Consistent with previous findings, maternal age was the strongest predictor of serum PCB levels, alongside year of sample collection, pre-pregnancy BMI, and polyunsaturated fatty acid levels. Twenty seven modules of placental DNA methylation were identified, including five which significantly correlated with one or more PCBs, and four which correlated with child neurodevelopment. Two modules associated with maternal PCB levels as well as child neurodevelopment, and mapped to *CSMD1* and *AUTS2*, genes previously implicated in ASD and identified as differentially methylated regions in mouse brain and placenta following gestational PCB exposure.

**Conclusions:** Placental DNA co-methylation modules were associated with maternal PCBs and child neurodevelopment. Methylation of *CSMD1* and *AUTS2* could potentially be mechanistically involved in ASD risk following maternal PCB exposure.

**GRAPHICAL ABSTRACT:** 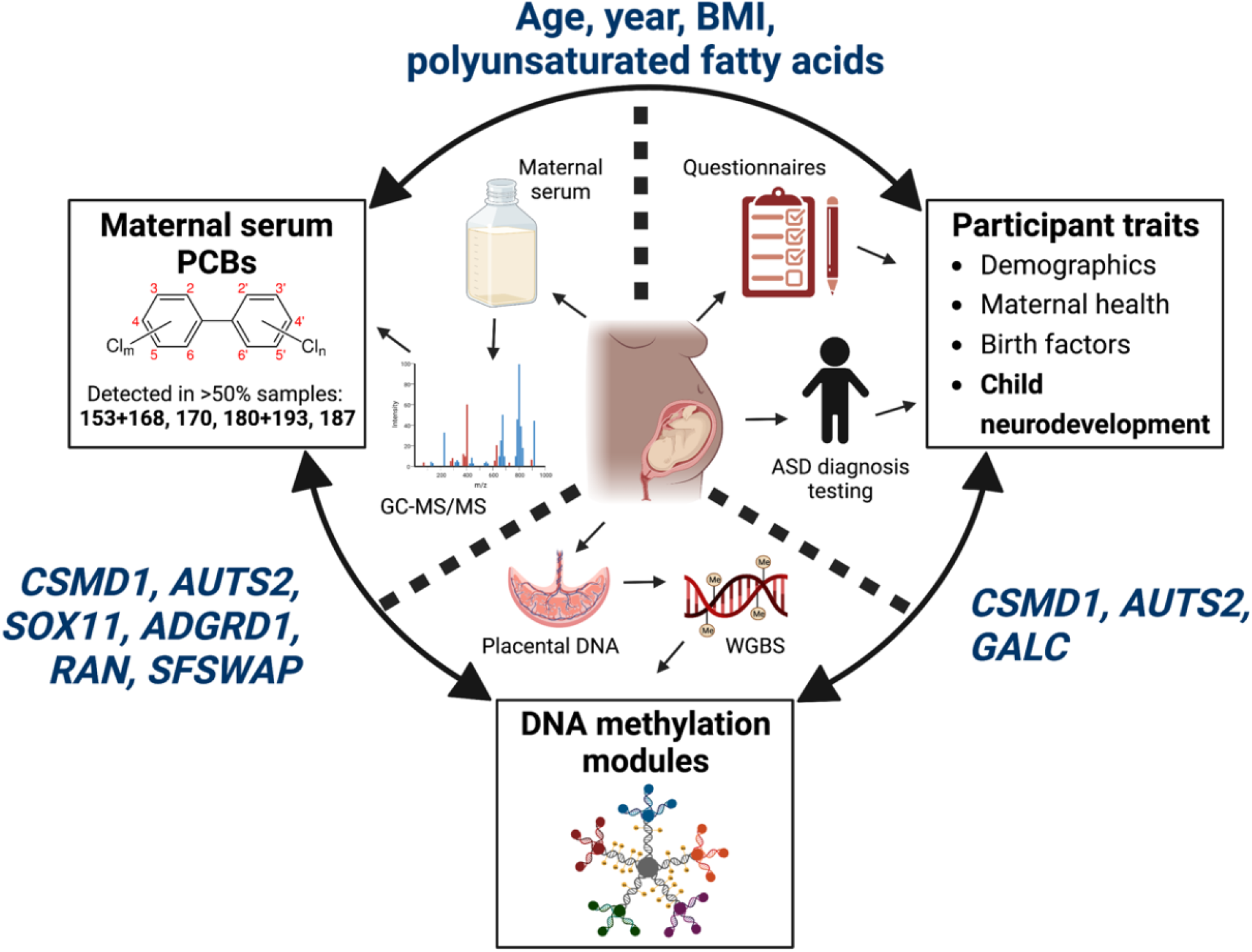

## 1. INTRODUCTION

Polychlorinated biphenyls (PCBs) are a class of 209 synthetic compounds that are ubiquitous in the environment and harmful to human health. Prior to their international ban in 2001 (Stockholm Convention, 2001), PCBs were commonly used in industrial processes and consumer products, such as plastics, paint, and pesticides. Over 20 years later, PCBs continue to be released into the environment from spills/improper disposal, degradation of PCB-containing materials, and inadvertent production (Grimm et al., 2015; Hu & Hornbuckle, 2010), resulting in their presence throughout our environment, from oceans (Wagner et al., 2019), soil (reviewed in (Wolska et al., 2014)) and landfills (reviewed in (Gabryszewska & Gworek, 2021), to human food products (Saktrakulkla et al., 2020) and indoor air, including air in schools (reviewed in (Herrick et al., 2016)). PCBs are detectable and persistent in many marine (reviewed in (Domingo & Bocio, 2007)) and land animal species (reviewed in (Rhind, 2002)), as well as in humans, including pregnant people (Granillo et al., 2019) and newborns (Berghuis et al., 2013; Mori et al., 2014; Yu et al., 2019). Adult exposure to PCBs by inhalation, absorption through skin, or consumption of contaminated foods such as dairy, meat and fish, increases risk for many diseases, including cancer (reviewed in (Leng et al., 2016; Wan et al., 2022)), cardiovascular disease (reviewed in (Gupta et al., 2018)) and reproductive abnormalities (reviewed in (Rattan et al., 2017)). Of significant concern, fetal exposure to PCBs via maternal blood has been associated with adverse health outcomes in the offspring, including increased risk of neurodevelopmental disorders such as autism spectrum disorder (ASD) (Panesar et al., 2020; Pessah et al., 2019).

ASD is a group of neurodevelopmental disorders characterized by impairments in social reciprocity and communication, restrictive interests, and repetitive behaviors. Risk for and protection from ASD is influenced by a wide variety of genetic and environmental factors as well as gene by environment interactions (reviewed in (Chaste & Leboyer, 2012; Cheroni et al., 2020; Lyall, Croen, Daniels, et al., 2017)). Diagnoses of ASD have been increasing over the past several decades and are currently estimated to affect approximately 1 in 44 children in the United States (Maenner, 2021). These sobering statistics are a primary premise for the hypothesis that gestational exposure to environmental chemicals influence individual risk for ASD. However, the mechanisms by which environmental factors influence risk remain controversial (Lein, 2015).

Epigenetic marks are widely posited as one mechanism mediating gene by environment interactions that confer ASD risk (Lein, 2015; Panesar et al., 2020) with the placental epigenome functioning as a potential biological substrate (Zhu et al., 2022). The placenta is a fetal tissue that invades the uterus and is responsible for providing nutrients and oxygen to the fetus as well as removing waste. While the placenta filters out many toxic chemicals from the maternal bloodstream, small lipophilic chemicals, such as PCBs, can cross the placental barrier and enter the bloodstream of the fetus (Guvenius et al., 2003; Park et al., 2008; Soechitram et al., 2004). Indeed, PCB levels in human placenta have been found to significantly correlate with those in maternal blood and umbilical cord blood (Jeong et al., 2018). This placental exposure can alter patterns of DNA methylation, a tissue-specific epigenetic mark that is influenced by both genetic and environmental factors. DNA methylation marks are dynamic during development when they are being established and then remain mostly stable, affecting gene expression throughout the lifetime (Moore et al., 2013).

Recently, differential DNA methylation has been identified in human placenta and umbilical cord blood of individuals later diagnosed with ASD (Mordaunt et al., 2020; Zhu et al., 2022). Moreover, maternal PCB levels have been associated with altered DNA methylation in human placenta (Ouidir et al., 2020) and in mouse placenta and fetal brain (Laufer et al., 2022). Both Ouidir et al. and Laufer et al. found enrichment for pathways related to neurodevelopment and neuronal/brain functions. However, no human study to date has examined associations between maternal PCB exposure, whole-epigenome placenta DNA methylation, and offspring neurodevelopmental outcomes.

In this study, we measured 209 PCB congeners from 104 samples of maternal serum collected at delivery and analyzed DNA methylation networks from 147 placental samples using weighted gene correlation network analysis (WGCNA) (Langfelder & Horvath, 2008) **(Figure 1)**. We correlated placental DNA methylation modules with maternal serum PCB values, as well as participant traits related to child neurodevelopment, maternal health, demographic factors, and birth factors. Using this approach, we identified known ASD risk genes *CSMD1* and *AUTS2* and other potential biomarkers of PCB exposure relevant to neurodevelopment.

**Figure 1.**
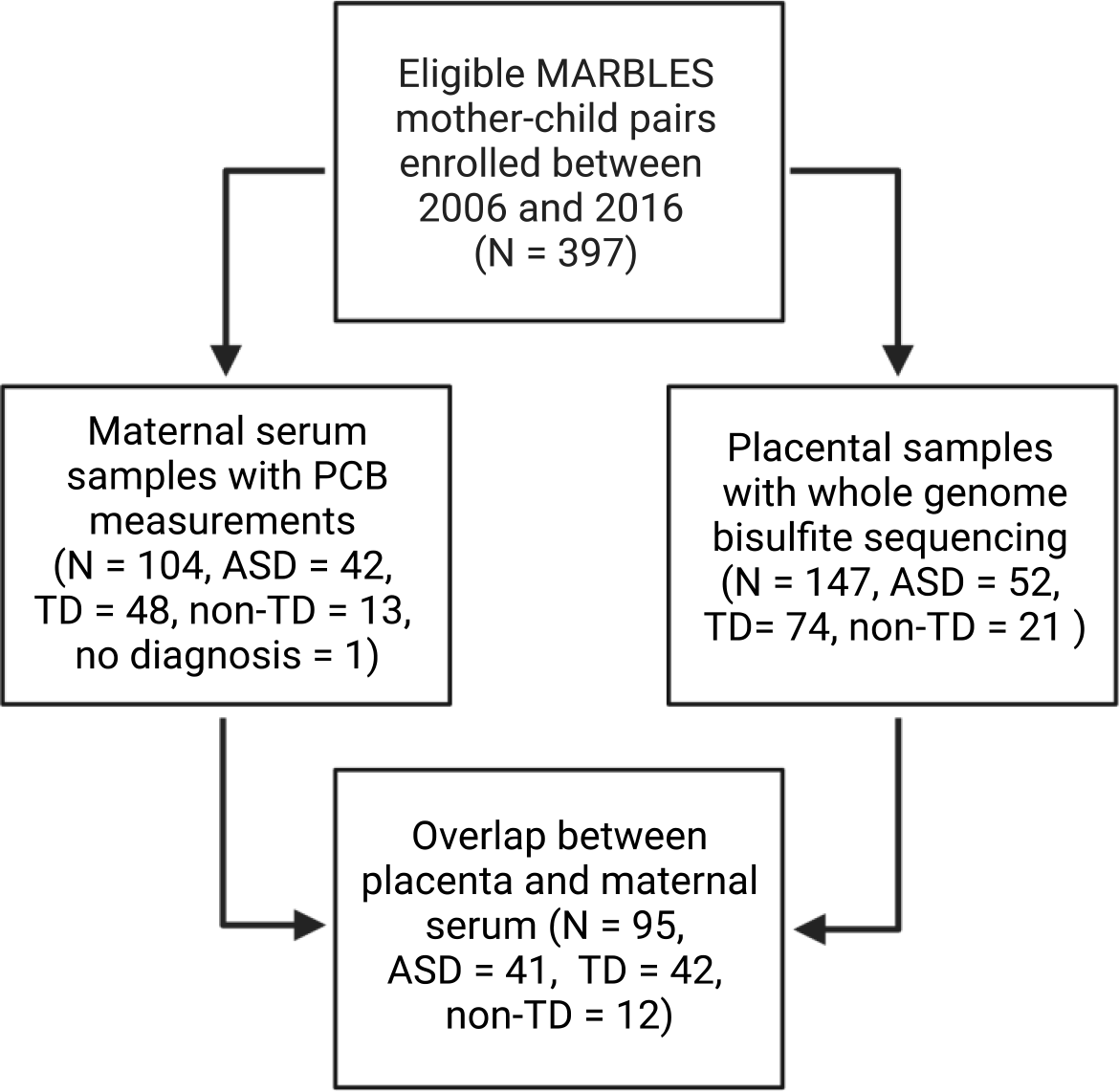
Sample inclusion from the MARBLES cohort

## 2. METHODS

### 2.1. Study Population

#### 2.1.1. MARBLES and EARLI cohorts

This study included mother-child pairs that were enrolled in the Markers of Autism Risk in Babies – Learning Early Signs (MARBLES) study from 2006-2016. As previously described (Hertz-Picciotto et al., 2018), MARBLES enrolls mothers in northern California who already have a biological child with ASD and are therefore at increased risk of delivering another child who will develop ASD. Enrollment in the MARBLES study required families to meet the following criteria: 1) the mother or father already had a biological child diagnosed with ASD, and the mother was 2) at least 18 years of age; 3) already pregnant or planning to become pregnant; 4) living within a 2-hour drive of the Davis/Sacramento area; and 5) proficient in English. This study used 104 maternal serum samples from MARLBES for PCB analysis and 147 placentas (after quality control) from MARBLES for DNA methylation analysis; 95 of the serum and placenta samples came from a mother-fetus pair.

This study uses an independent replication cohort from the Early Autism Risk Longitudinal Investigation (EARLI) to validate results from DNA methylation analysis. Similarly to MARBLES and previously described elsewhere (Newschaffer et al., 2012), EARLI enrolls mothers who already have a biological child with ASD. Families were recruited from Drexel/Children’s Hospital of Philadelphia, Johns Hopkins/Kennedy Krieger Institute, Kaiser Permanente Northern California, and University of California, Davis. Enrollment in the EARLI study required families to meet the following criteria: 1) the mother or father already had a biological child diagnosed with ASD, and the mother was 2) at least 18 years of age; 3) less than 29 weeks pregnant; 4) living within a 2-hour drive of the study site; and 5) proficient in English or Spanish. This study used 47 placentas from EARLI to confirm the preservation and quality of the DNA co-methylation modules.

#### 2.1.2. Diagnostic classification for autism spectrum disorder

Children enrolled in MARBLES and EARLI were clinically assessed for autism spectrum disorder (ASD) around 36 months of age by professional examiners. Diagnosis was based on the Autism Diagnostic Observation Schedule (ADOS) (Lord et al., 2000), Autism Diagnostic Interview – Revised (ADI-R) (Lord et al., 1994), and Mullen Scales of Early Learning (MSEL) (Mullen, 1995). ADOS is a standardized assessment tool that uses structured and semi-structured activities that allow the examiner to evaluate communication skills, social interaction, and imaginative use of materials. The ADOS module 2 scores range from 0 to 28, with scores equal to or higher than 7 meeting the cut-off for ASD. ADI-R is a structured interview with a parent or caregiver to discuss the developmental history and current behavior of the child; this interview can be useful to distinguish ASD from other developmental disorders. MSEL is a standardized evaluation whereby the child completes a set of motor, visual, and language tasks which are assessed by the examiner. MSEL has four subscale scores (fine motor, expressive language, receptive language, and visual reception) as well as an overall early learning composite score, which is referred to as Mullen Score in this manuscript. The composite score range is 49 to 155, with scores below 70 indicating neurodevelopmental delays.

Based on ADOS and MSEL scores, children in MARBLES and EARLI were categorized into three outcome groups: ASD, typical development (TD), and non-typical development (non-TD)(Chawarska et al., 2014; Ozonoff et al., 2014; Schmidt et al., 2019). Children classified in the ASD group had ADOS scores above the cut-off and met the *DSM-5* criteria for ASD. Children classified in the TD group had MSEL scores that were all within 2 SD and no more than one MSEL subscale score that was 1.5 SD below the normative mean, as well as ADOS scores that were 3 or more points below the cut-off score for ASD. Children that met the criteria for neither ASD nor TD were classified as non-TD; these children had low MSEL scores (two or more MSEL subscale scores more than 1.5 SD below the normative mean or at least one MSEL subscale score more than 2 SD below the normative mean) and/or high ADOS scores (within 3 points of the cut-off score for ASD).

#### 2.1.3. Participant traits

MARBLES participants provided demographic, nutritional, environmental, pregnancy, and medical information through telephone-assisted interviews and mailed questionnaires. We selected particular traits of interest, coded them numerically for analysis, and grouped them into four categories for ease of discussion: demographics, maternal health, birth factors, and child neurodevelopment. Information on some traits was not available for all participants, in which case the data were removed and statistical analyses performed on the data available.

Demographic traits evaluated include both parents’ birthplace (California/not California), race (white/non-white), ethnicity (Hispanic/non-Hispanic), education level (less than high school/high school diploma or GED/some college/Bachelor’s degree/graduate or professional degree), and age at time of the child’s birth, as well as home ownership (renter/owner), delivery payer (public/private insurance at time of child delivery), and sex assigned at birth of the enrolled child (male/female). Maternal health factors evaluated include diagnosis of gestational diabetes (yes/no), pre-eclampsia (yes/no), or chronic hypertension (yes/no), as well as maternal BMI pre-pregnancy (continuous; calculated from weight and height), maternal plasma polyunsaturated fatty acid (PUFA) levels (DPA + DHA + EPA measured in maternal serum), maternal omega 3s (high/low), prenatal vitamin use in month 1 of pregnancy (yes/no), and maternal smoking during pregnancy (yes/no). PUFA and omega 3 data have been previously reported (Huang et al., 2020). Birth factors evaluated include delivery method (vaginal/C-section), birth month, birth season (fall, winter, spring, summer), birth year (2006-2016), parity (number of pregnancies over 20 weeks gestation prior to this pregnancy), Apgar score at 5 minutes, Apgar score at 1 minute, birth weight (grams), birth length (cm), and gestational age at delivery (number of weeks into pregnancy when delivery occurs). Factors related to child neurodevelopment include age at which ADOS and MSEL tests were performed, MSEL composite score, ADOS comparison score, and diagnosis of ASD (ASD, TD, non-TD).

Participant traits were correlated with one another using Pearson and Spearman correlations and *p*-values were corrected for multiple testing by FDR using the psych R package v2.2.5 corr.test function (Revelle, 2022). Pearson measures linear correlation, whereas Spearman measures monotonic correlation, providing less restrictive but also less powerful results. Given the structural diversity of our data, we elected to use both correlation measures to capture as many potential relationships as possible. We adjusted *p*-values by FDR due to its power to detect true positives (Benjamini & Hochberg, 1995, 2000; Chen et al., 2010; Korthauer et al., 2019). GraphPad Prism version 9.4.1, GraphPad Software, San Diego, California USA, www.graphpad.com, chi-squared test was used to test if prenatal vitamin use differed among ASD, non-TD, and TD diagnostic groups, while one-way ANOVA and Tukey’s multiple comparisons test were used to test differences in MSEL scores across diagnostic groups.

### 2.2. PCB measurements and analysis

#### 2.2.1. Serum collection and storage

Maternal serum was collected in red-top serum-separator vacutainers at time of delivery and processed with centrifugation within 4 hours if possible, and stored frozen at -80 degrees Fahrenheit. For this analysis, specimen aliquots were selected based on: having a minimum of 6 mL at delivery, child diagnosis data available, and a placenta specimen available for WGBS. Serum aliquots were de-identified and shipped to the Analytical Core of the Iowa Superfund Research Program frozen on dry ice. A pooled serum specimen from samples with ample volume was created and included in each analysis run for quality assurance purposes. In this paper, PCB congeners are referred to by their Ballschmitter and Zell number (US EPA, 2015).

#### 2.2.2. Chemicals and materials

^13^C-labeled PCBs, including ^13^C_12_-PCB 3, ^13^C_12_-PCB 15, ^13^C_12_-PCB 31, ^13^C_12_-PCB 52, ^13^C_12_-PCB 118, ^13^C_12_-PCB 153, ^13^C_12_-PCB 180, ^13^C_12_-PCB 194, ^13^C_12_-PCB 206, and ^13^C_12_-PCB 209, were purchased from Cambridge Isotope Laboratories Inc. (Andover, Massachusetts, USA). Deuterated PCB 30 (d_5_-PCB 30) was obtained from C/D/N Isotopes (Pointe-Claire, Quebec, Canada). A calibration standard containing all 209 PCB congeners and PCB 204 (internal standard) were purchased from AccuStandard, Inc. (New Haven, Connecticut, USA). Unique chemical identifiers of all 209 PCB congeners have been reported previously (Li, Westra, et al., 2022). Pesticide grade solvents, such as hexane and dichloromethane, were purchased from Fisher Scientific (Pittsburgh, Pennsylvania, USA). The Standard Reference Material (SRM 1957) was obtained from the National Institute of Standards and Technology (NIST, Gaithersburg, Maryland, USA).

#### 2.2.3. Extraction of PCBs from maternal serum

Serum samples were extracted and analyzed by gas chromatography-tandem mass spectrometry (GC-MS/MS) analysis using published procedures (Li, Hefti, et al., 2022; Marek et al., 2014; Milanowski et al., 2010) in the Analytical Core of the Iowa Superfund Research Program. Briefly, tissue samples were homogenized in 3 mL isopropanol. One mL of diethyl ether was added, and samples were spiked with isotope-labeled PCB (5 ng/each in hexane). The tubes were inverted for 5 minutes and centrifuged at 1,378 g for 5 minutes. The organic layer was transferred to a new tube. The residue was re-extracted with 1 mL of isopropanol and 2.5 mL of hexane and diethyl ether (9:1, vol/vol). The combined organic extracts were washed with 5 mL of phosphoric acid (0.1 M in 0.9% aqueous sodium chloride), and the aqueous phase was re-extracted with 1 mL of hexane and diethyl ether (9:1, v/v). The combined extracts were concentrated to near dryness under a gentle stream of nitrogen, reconstituted in 4 mL of hexane, and 2 mL of a potassium hydroxide solution (0.5 M in water-ethanol, 1:1, v/v) was added. After inversion for 5 min and centrifugation at 1,378 *g* for 3 min, the organic phase was separated, and the bottom aqueous phase was re-extracted with 3 mL of hexane. The combined organic phase contained the PCBs. The PCB fraction was concentrated to ∼ 0.5 mL under a gentle stream of nitrogen and passed through a glass cartridge filled with 2 g of acidified silica gel (silica gel and concentrated sulfuric acid, 2:1, w/w) with 0.2 g activated silica gel at the bottom and prewashed with 3 mL of hexane. PCBs were eluted from the cartridge with 14 mL of hexane. The eluent was concentrated to 4 mL and treated with 2 mL of concentrated sulfuric acid. After inversion for 5 min and centrifugation at 1,378 g for 3 min, the organic phase was collected, and the sulfuric acid layer was re-extracted with 3 mL of hexane. The combined organic extract was concentrated to about 50 µL and transferred to a glass autosampler vial with an insert. Before analysis, the sample was spiked with the internal standards (d5-PCB 30 and PCB 204; 5 ng each in hexane).

#### 2.2.4. GC-MS/MS determination of PCBs

PCB samples were analyzed on an Agilent GC system coupled with an Agilent 7000 Triple Quad in the multiple reaction monitoring (MRM) mode on an SPB-Octyl capillary column (30 m length, 250 µm inner diameter, 0.25 µm film thickness; Sigma-Aldrich, St. Louis, Missouri, USA). The following temperature program was used: 45 °C, hold for 2 min, 100 °C/min to 75 °C, hold for 5 min, 15 °C/min to 150 °C, hold for 1 min, 2.5 °C/min to 280 °C, and hold for 5 min. The injector temperature program is as follows: start at 45 °C, hold for 0.06 min, 600 °C/min to 325 °C and hold for 5 min. The transfer line temperature was 280 °C. Helium was used as a carrier gas with a constant flow rate of 0.8 mL/min. The precursor-product ion transitions of all PCB analytes used for the MS/MS analysis have been reported previously (Li, Hefti, et al., 2022).

#### 2.2.5. Quality assurance/quality control for the GC-MS/MS analysis

Extraction efficiency, reproducibility, and accuracy were assessed using ^13^C-labeled surrogate standards, method blanks, and analysis of a standard reference material (SRM 1957, NIST). Limits of quantification (LOQs) were calculated as 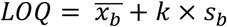, where 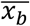 indicates the mean of method blank results, *k* indicates the Students′ *t*-value appropriate for the single-tailed 99^th^ percentile *t* statistic and a standard deviation estimate with n-1 degree of freedom, and *s*_*b*_ indicates the sample standard deviation of the replicated method blank samples. The recovery rates of the surrogate standards ranged from 70±10 % to 97±9 %.

#### 2.2.6. Statistical analysis of PCBs and PCB-participant trait relationships

PCB congeners were assessed for detection frequency, defined as the percent of the 104 materal serum samples with PCB values over the LOQ, and labeled as detected (non-censored) or non-detected (censored) for each sample. Individual or co-eluting congeners that were detected in ≥50% (n=52) of samples were selected for further analysis due to the increased robustness of statistical methods. PCBs 153+168, 170, 180+193, and 187 met this cutoff. All PCB congener values (ng) were then adjusted by maternal serum weight (g), and these serum weight-adjusted values were used in all remaining analyses. PCB analyses described in this section were completed with the NADA R Package v1.6-1.1 using R v4.1.3, unless otherwise specified (Helsel, 2005). NADA is optimized for censored environmental data (Lee, 2020)

Summary statistics (mean, median, standard deviation) were estimated for PCBs 153+168, 170, 180+193, and 187 using the censtats function, which reports statistics from three methods: reverse Kaplan-Meier (a non-parametric test that does not assume a normal distribution of the data), Robust Regression on Order Statistics (ROS; a semi-parametric model that assumes the detected data is normally distributed but makes no assumptions about the censored data), and Maximum Likelihood Estimation (MLE; a parametric model that assumes a normal distribution of all data). The data were log-transformed for ROS and MLE to attain normality, due to the right-skew of the non-transformed data. All three models avoid removal, replacement, or imputation of the nondetected values, instead estimating statistics using all data points.

PCBs 153+168, 170, 180+193, and 187 were correlated with one another using Kendall’s tau for left-censored data, a nonparametric correlation coefficient that measures the monotonic association between two variables, one or both of which may include censored data. Both detected and non-detected data were included in the correlations. MARBLES participant traits (related to demographics, maternal health, birth factors, and child neurodevelopment) were correlated with PCBs 153+168, 170, 180+193, and 187 values using Kendall’s tau for left-censored data, and *p*-values were corrected by FDR. To assess which participant traits most contributed to the levels of PCBs in the maternal serum, we computed fitted multiple linear regression models. First, trait-PCB congener pairs were filtered to those that were associated with a raw *p*-value < 0.2. If multiple trait-PCB congener pairs met this cut-off and the traits were correlated with each other, r > 0.5, the trait-PCB pair with the less significant *p*-value was removed from the analysis. This was done to remove multicollinearity and increase the reliability of the model. With the remaining PCB-trait pairs, a fitted multiple linear regression equation for censored data was computed using the cenmle function. We then assessed detection frequencies and congener levels for all congeners across maternal age groups (>35 vs ≤35), offspring birth year (2006-2011 vs 2012-2016), maternal BMI (>25 vs ≤25), offspring ASD diagnosis (ASD vs non-TD vs TD), and offspring sex (male vs female) using coin R package v1.4.2, VGAM R package v1.1.6, and censReg R package v0.5.34.

To further examine the relationship between maternal serum PCB levels and diagnosis of ASD in the offspring, the Peto-Prentice non-parametric test was used to test the means of PCB values between diagnostic groups (ASD/non-TD/TD). The Peto-Prentice test reports two-sided *p*-values which were divided in two to produce one-sided *p*-values. To further examine the relationship between maternal serum PCB levels and maternal pre-pregnancy BMI, we correlated PCB-BMI tau with K_ow,_ the octanol-water partition coefficient, for each PCB: 153+168, 170, 180+193, and 187 (Hawker & Connell, 1988). For co-eluting PCB congeners (PCBs 153+168 and 180+193), we averaged the K_ow_ of the two congeners.

### 2.3. Placental DNA methylation analysis

#### 2.3.1. Whole genome bisulfite sequencing (WGBS)

The placental samples and WGBS data used in this study were previously described (Ladd-Acosta et al., 2021; Zhu et al., 2022). Briefly, 157 MARBLES and 47 EARLI placental samples were frozen within 4 hours of birth. DNA was extracted from the placental samples and treated with sodium bisulfite. Following whole genome sequencing of bisulfite-treated DNA, sequencing files were processed, aligned to the hg38 genome, and converted to CpG methylation count matrices.

#### 2.3.2. Placental DNA co-methylation network analysis

We analyzed placental DNA methylation using Comethyl, v1.1.1 with R v 4.1.0, a published R package that performs weighted gene correlation network analysis (WGCNA) for whole-genome bisulfite sequencing (WGBS) data (Mordaunt et al., 2022). Comethyl forms modules from groups of genomic regions that have correlated methylation, meaning that the proportion of CpGs that are methylated in a given region correlate with the proportion of CpGs methylated in other regions across samples. Modules are biologically informative because regions with correlated methylation tend to be part of the same biological pathways and are less susceptible to random variation s in methylation than are individual CpG sites. In order to build modules from genomic regions that are particularly informative, we filter regions to include only those that have variable methylation across samples and therefore, may contribute to differential phenotypes. Methylation of a given module is represented by a single value for each sample, called the “eigennode”, where a more positive eigennode value reflects higher methylation levels and a more negative value reflects lower methylation.

Specifically, 157 individual placenta Bismark CpG reports were read into a single BSseq object that was used to calculate the number of CpGs at different filtering options for both sequencing coverage and number of samples. Ten samples were removed due to low sequencing coverage, resulting in 147 samples being used for analysis. CpGs with at least 4x coverage in at least 80% of the 147 samples were selected to form regions, resulting in 13,056,694 filtered CpGs (44% of CpGs). Regions, defined as 3 or more CpGs separated by no more than 150 base pairs, were filtered for those with at least 12x coverage and a methylation standard deviation of at least 5%, resulting in 199,944 filtered regions. The methylation data was then adjusted for the top 26 principal components to adjust for potential confounding variables (Parsana et al., 2019). Using the adjusted methylation data, scale-free topology with Pearson correlations was assessed to evaluate fit and connectivity of the model. A soft power threshold of 15 was selected, meaning that all correlations were raised to the power of 15 to increase the strength of strong correlations and minimize weak correlations and background noise. Finally, modules were identified using a two-phase clustering approach that reduces computational intensity: filtered regions were grouped into 6 blocks using k-means clustering, then a full network analysis was performed for each block. Modules were identified in each block through hierarchical clustering and adaptive branch pruning, then were merged if their eigennode values were highly correlated (r > 0.9). Eigennode values are a single value that summarizes the methylation of a module for each sample. Regions not assigned to a module were assigned to the grey module.

#### 2.3.3. Co-methylation module, sample, and trait correlations

Once modules were identified, we correlated module eigennode values with one another to identify highly related pairs. Correlations were calculated by Bicor, and *p*-values were derived based on Fisher’s transformation.

We then correlated module eigennode values with MARBLES participant traits (related to demographics, maternal health, birth factors, and child neurodevelopment) as well as measured maternal serum PCB values. Module eigennode values were correlated with participant traits using Bicor, and with PCB values using Kendall’s tau for left-censored data (NADA R package).

#### 2.3.4. Co-methylation module annotation

Module regions were annotated to genes using the Genomic Regions Enrichment of Annotations Tool (GREAT) and the hg38 genome (McLean et al., 2010). A region assigned to a module was annotated to a gene if its genomic coordinates fell within the gene’s regulatory domain, defined as 5 kb upstream and 1 kb downstream from its transcription start site, or up to 1 Mb from the transcription start site until the nearest gene’s regulatory domain. Regions can map to zero, one, or multiple genes. Gene information was provided by BioMart while gene context and CpG context were provided by annotatr (Cavalcante & Sartor, 2017). Genic features include promoters (<1kb upstream of a transcription start site), 5’UTRs, exons, introns, 3’UTRs, enhancers, intergenic regions, and regions 1kb-5kb upstream of a transcription start site (Cavalcante & Sartor, 2017). CpG features include CpG islands (regions of the genome with a high frequency of CpG sites as defined by UCSC Golden Path), CpG shores (the 2kb upstream and downstream of CpG island boundaries), CpG shelves (the 2kb upstream and downstream of CpG shores), and the open sea (beyond 4kb from CpG island boundaries) (Cavalcante & Sartor, 2017).

We analyzed the overlap between module regions and partially methylated domains (PMDs) using placenta PMDs previously identified by a machine learning approach on the hg18 genome (Schroeder et al., 2013). We used the UCSC liftover tool to lift the hg18 PMDs to hg38, then compared regions assigned to modules to PMDs; a region was considered within a PMD if the entire region fit in the PMD.

#### 2.3.5. Co-methylation module preservation

To assess the consistency of modules generated by the MARBLES samples we performed module preservation using an independent dataset from the EARLI study.We called modules from the EARLI dataset using CpGs and regions restricted to those covered in the MARBLES dataset. Methylation data was corrected for the top 16 principal components, and a soft power threshold of 23 was selected to achieve a high fit for the model. Pearson correlation was then used to identify associations between regions and to call modules. We evaluated the MARBLES and EARLI modules through multiple statistics, assessing the quality, preservation, separability, and accuracy of each module (Langfelder et al., 2011). Module assignments are permuted 100 times to calculate Z-scores and P-values. Based on simulation studies by Langfelder et al., 2011, we considered Z-scores over 10 to be strong evidence for module preservation and Z-scores between 2 and 10 to be weak-moderate evidence.

## 3. RESULTS and DISCUSSION

### 3.1. Highly chlorinated PCB congeners are most frequently detected in maternal serum

We measured levels of all 209 PCB congeners in 104 MARBLES study samples of maternal serum collected at time of delivery **(Figure 2A) (Supplemental Tables S1 and S2)** (Hertz-Picciotto et al., 2018). While most human PCB studies use serum collected during pregnancy, delivery serum provides a unique timepoint to study lipid-soluble PCBs because maternal plasma free fatty acids have been found to double during labor (Kashyap et al., 1976). In 27 instances, two-four PCB congeners co-eluted, resulting in 173 PCB measurements per sample. Each sample had between zero and 41 PCB congeners detected, with a mean of 6.68 and a median of 4 congeners detected per sample. Eighty-one single or co-eluting PCB congeners were detected in zero samples; 76 in 1-10% of samples; 5 in 11-20% of samples; 7 in 21-50% of samples; and 4 in ≥50% of samples **(Supplemental Table S1)**. These detection frequencies are lower than those in postmortem human brain using similar methods (Li, Hefti, et al., 2022). In both studies, highly chlorinated PCBs were more likely to be detected than lower chlorinated PCBs, despite the decreased response of the instrument as the degree of chlorination increases. This may be explained by highly chlorinated PCB congeners’ resistance to metabolization, leading to bioaccumulation in the body (Kato et al., 1980; Mathews & Anderson, 1975; Mills et al., 1985).

**Figure 2.**
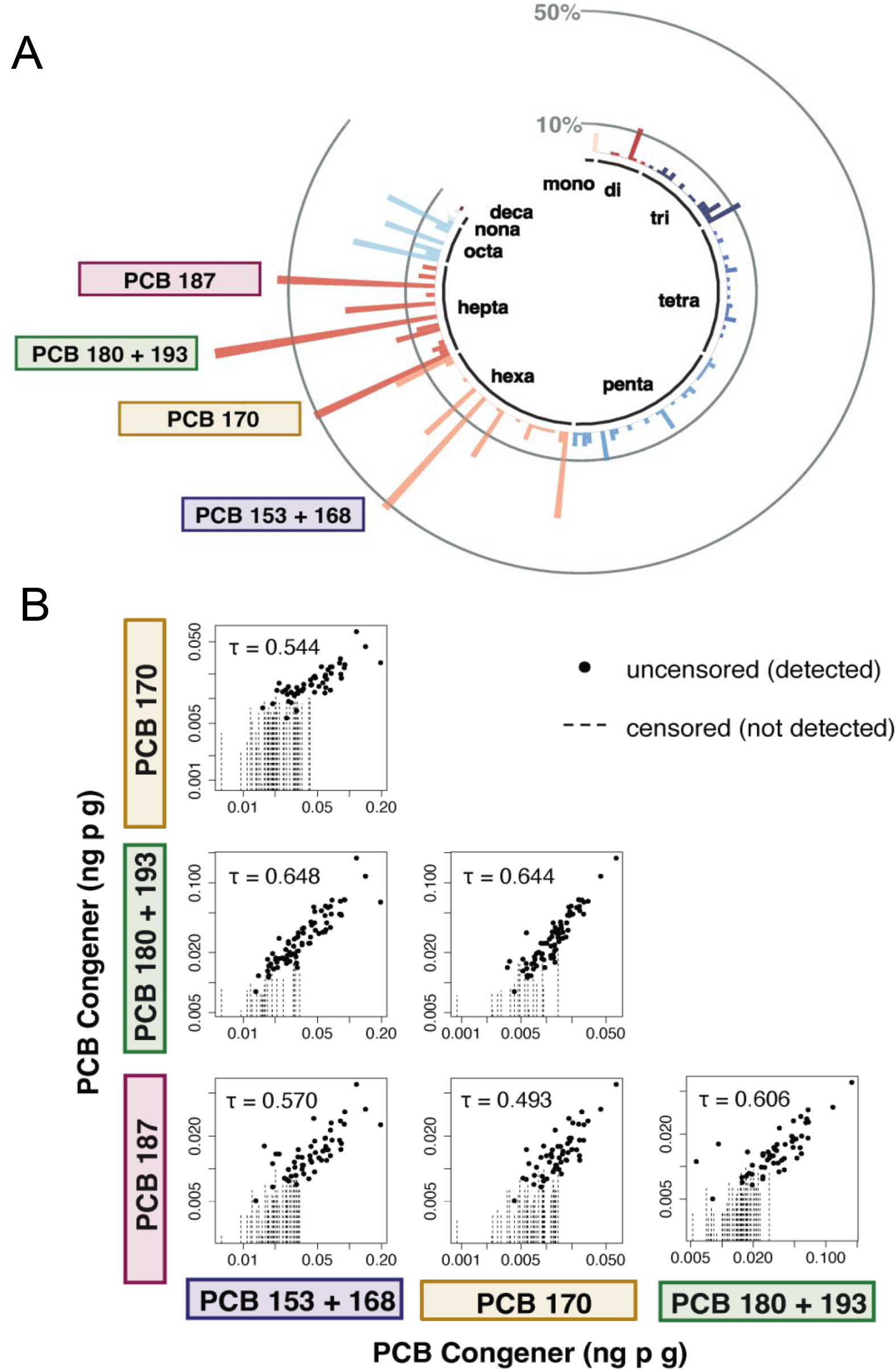
The most highly detected PCB congeners are 153 + 168, 170, 180 + 193, and 187, all of which are highly correlated. **A)** Detection frequencies of PCB congeners in maternal delivery serum samples. PCB congeners are plotted in the order of their Ballschmiter and Zell number, as defined by the EPA (US EPA, 2015). Grey lines indicate 10% and 50% detection frequencies and black labels indicate the number of chlorines. The PCB congeners detected in over 50% of samples are labelled by name. **B)** Correlation plots for PCBs 153 + 168, 170, 180 + 193, and 187 where dots represent uncensored data and dashed lines represent censored data. Kendall’s tau correlation coefficient for each comparison is represented by τ

The four PCB congeners/congener pairs that were detected in ≥50% (n=52) of samples were selected for further analysis due to the increased robustness of statistical methods when censored data points represent less than half the data: PCBs 153s + 168, 170, 180 + 193, and 187. These PCB congeners were also detected in ≥50% of postmortem brain samples (Li, Hefti, et al., 2022) and found at high concentrations in maternal blood plasma, cord blood plasma, breast milk (Guvenius et al., 2003), and placenta (Naqvi et al., 2018). No PCB congeners were detected in fewer than 50% of all samples but in greater than 50% of one of the ASD diagnostic groups (ASD/non-TD/TD), indicating that we did not miss a congener that may discriminate between diagnostic groups.

### 3.2. Highly detected PCB congeners (153 + 168, 170, 180 + 193, and 187) have variable but correlated levels in maternal serum

We estimated summary statistics (mean, median, standard deviation) for PCBs 153 + 168, 170, 180 + 193, and 187 **(Table 1 and Supplemental Table S3)** using both the detected and non-detected data in the models, which has been shown to be more effective than substituting or replacing non-detected data points (Ahmadi et al., 2021; Canales et al., 2018). Inputted PCB values were adjusted by serum weight, which did not differ significantly across ASD diagnostic groups (ASD/non-TD/TD) **(Supplemental Figure S4)**. PCBs 153 + 168 and 180 + 193 had higher median and mean concentrations than PCBs 170 or 187. PCB 153 and PCB 180 have previously been detected as among the 12 most abundant PCB congeners in maternal serum in the MARBLES cohort (Sethi et al., 2019), as well as highly detected in brain tissue (Chu et al., 2003; Corrigan et al., 1998; Dewailly et al., 1999; Li, Hefti, et al., 2022; Mitchell et al., 2012). Additionally, PCB 153 + 168 are the least lipophilic of this group, meaning they are more readily released from adipose tissue into the blood, potentially resulting in higher average serum levels (Louis et al., 2016).

**Table 1.** Estimated summary statistics for PCB congeners in maternal serum by Regression on Order Statistics.

| PCB Congener | Detected (n,%) | Median (ng/g) | Mean (ng/g) | SD (ng/g) |
| --- | --- | --- | --- | --- |
| PCB 153 + 168 | 54 (51.9) | 0.0283 | 0.0368 | 0.0301 |
| PCB 170 | 53 (51.0) | 0.0069 | 0.0114 | 0.0091 |
| PCB 180 + 193 | 80 (76.9) | 0.0192 | 0.0272 | 0.0240 |
| PCB 187 | 56 (53.8) | 0.0078 | 0.0104 | 0.0086 |

We next tested the hypothesis that measured values of PCB congeners in serum correlate with one another. Confirming previous findings (DeVoto et al., 1997; Pauwels et al., 1999), PCBs 153 + 168, 170, 180 + 193, and 187 were all significantly correlated (tau correlation coefficients ranged from 0.48 to 0.63, with all *p*-values < 10^−12^), though correlation coefficients were lower than in previous studies, likely due to our inclusion of non-detected data points **(Figure 2B) (Supplemental Table S4)**.

### 3.3. Participant traits related to demographics, maternal health, birth factors, and child neurodevelopment are correlated

Next, we took advantage of the large volume of metadata collected within the MARBLES study to explore relationships between traits related to demographics, maternal health, birth factors, and child neurodevelopment **(Supplemental Tables S5 and S6)**. To identify linear and monotonic relationships, we correlated all numerically-coded participant traits using Pearson (linear) **(Figure 3A) (Supplemental Tables S7, S9)** and Spearman (monotonic) **(Supplemental Figure S2) (Supplemental Tables S8, S10)** correlations. Spearman detected 54 significant trait-trait correlations (FDR < 0.05), two of which were not detected by Pearson, while Pearson detected 55, three of which were not detected by Spearman, indicating the value of performing both methods. Unsurprisingly, traits within a given category were often strongly correlated, including demographic traits of parental ages, parental education levels, home ownership, and delivery payer (public or private insurance), as well as paired demographic traits of the two parents such as birthplace, race, ethnicity, age, and education. Birth factors, including birth weight and birth length, as well as maternal health factors, including omega 3 and plasma PUFA levels, were also strongly correlated (FDR < 0.05).

**Figure 3.**
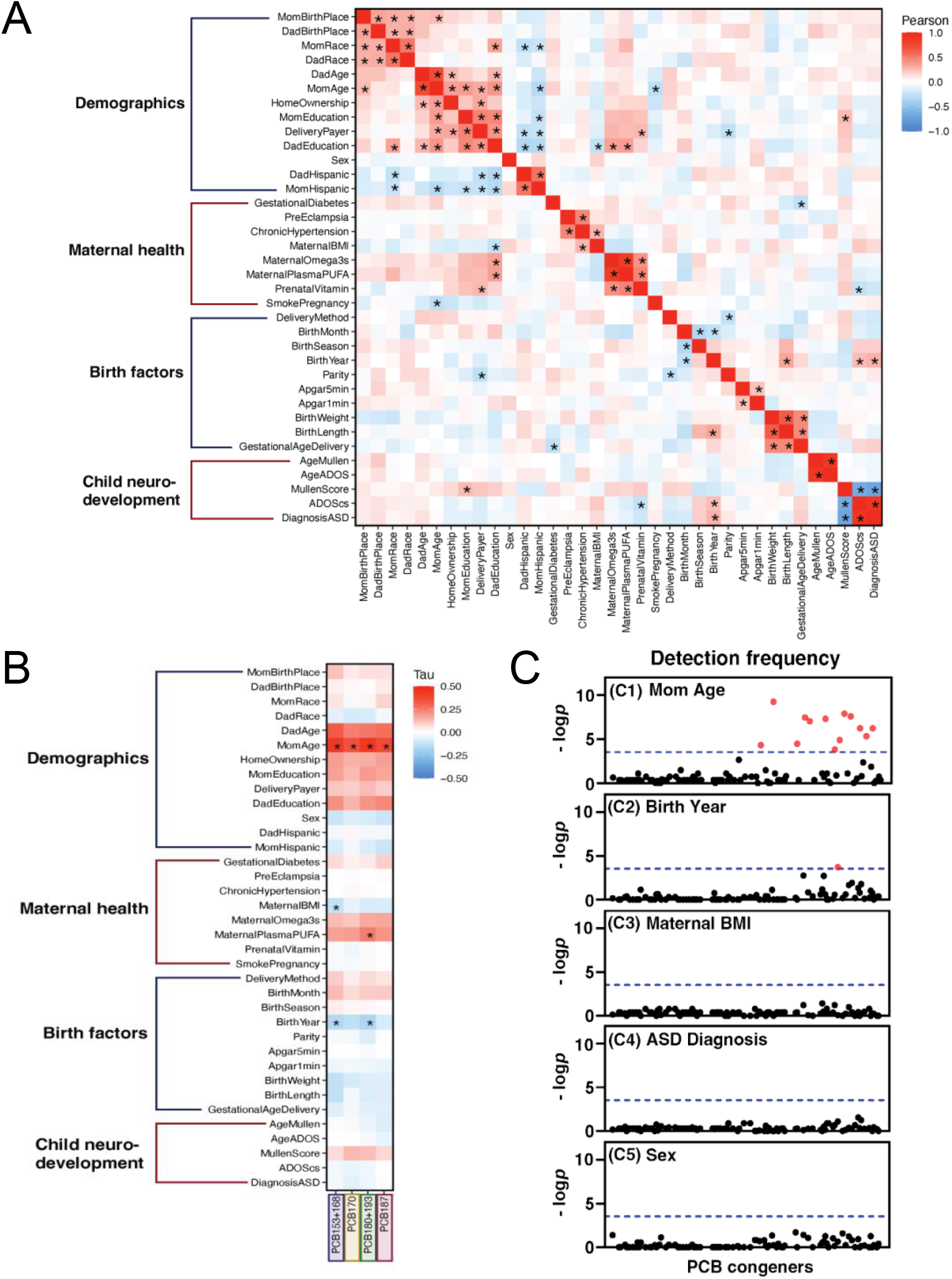
Participant traits correlate with one another and with maternal serum PCBs. **A)** Trait-trait correlations with Pearson correlation. Traits are grouped into four main categories and ordered hierarchically within each category. *FDR < 0.05. **B)** Participant traits correlated with maternal serum PCB concentrations using Kendall’s tau for left-censored data. * indicates significance in the multiple linear regression model for a given PCB congener/congener pair. **C)** Plots of *p*-values comparing detection frequencies of single or co-eluting PCB congeners between mom age groups (>35 vs ≤35), offspring birth year (2006-2011 vs 2012-2016), maternal BMI (>25 vs ≤25), offspring ASD diagnosis (ASD vs non-TD vs TD), and offspring sex (male vs female). The dotted line indicates the value of log *p* x (−1) of Bonferroni-adjusted multiple comparisons (*p* = 2.89 × 10^−4^) (Li et al., 2022). Detection frequencies were determined in 104 serum samples for 209 PCB congeners, analyzed as 173 peaks of single or co-eluting congeners. Red dots in panels (C1 and C2) indicate PCB congeners with a significant difference. PCB congeners are plotted on the x-axis in the order of their Ballschmiter and Zell number, as defined by the EPA (US EPA, 2015). Congeners with *p* = 1 were not plotted. The *p* values of the corresponding PCB congeners are presented in Table S17.

ASD diagnosis strongly correlated with ADOS comparison score (FDR = 0) and Mullen score (FDR = 0), which are both used to diagnose ASD, as well as birth year (FDR < 0.01), with rates of diagnosis increasing from 2006 to 2016. This aligns with previous findings that ASD is increasing in prevalence and/or diagnostic rates (CDC, 2022). ADOS comparison score correlated (FDR < 0.05) with prenatal vitamin use, which has previously been shown to be protective for ASD (Levine et al., 2018; Schmidt et al., 2011, 2019). To further examine this relationship in our cohort, we used chi-squared to test if prenatal vitamin use in month 1 of pregnancy (yes/no) was significantly different for ASD diagnostic groups (TD/non-TD/ASD). Aligning with previous findings, prenatal vitamin use was associated with ASD diagnosis in the protective direction (*P* = 0.0062) **(Supplemental Figure S3)**. Additionally, Mullen Score was significantly associated with mom education (FDR = 0.02), with greater educational attainment as child Mullen Score increased. Higher Mullen scores indicate greater cognitive development and decreased likelihood of ASD diagnosis **(Supplementary Figure S4)**. Low maternal education has been associated with increased risk and severity of ASD in previous studies (Dong et al., 2022; Leonard et al., 2005; Schoultz et al., 2010).

### 3.4. PCB levels in maternal serum are correlated with participant traits, particularly maternal age, but not child ASD diagnosis

Next, we explored associations between participant traits and levels of PCBs 153 + 168, 170, 180 + 193, and 187 in maternal serum collected at delivery **(Figure 3B)**. Based on existing literature, we hypothesized that maternal serum PCB levels would be positively associated with diagnosis of ASD in the offspring (Bernardo et al., 2019; Lyall, Croen, Sjödin, et al., 2017), negatively associated with maternal BMI (Koh et al., 2016; Lan et al., 2021; Mullerova et al., 2008), and negatively associated with child birth year (year of maternal serum sample collection), with the latter prediction reflecting data indicating that environmental levels of highly chlorinated PCBs, including PCBs 153 + 168, 170, 180 + 193, and 187, are decreasing over time (Brändli et al., 2007; Jones et al., 1992).

Kendall’s tau for left-censored data was estimated for each PCB-trait pair, and *p*-values were adjusted by FDR **(Supplemental Table S11, S12, S13)**. PCBs 153 + 168, 170, 180 + 193, and 187 were not significantly (FDR < 0.05) associated with ASD diagnosis, which we confirmed by the Peto-Prentice test **(Supplemental Table S14)**. At FDR < 0.05, PCBs 153+168 and 187 were significantly associated with maternal and paternal ages, paternal education level, and home ownership (renter/owner); PCB 170 with maternal and paternal ages; and PCB 180+193 with maternal and paternal ages and education levels **(Supplemental Figure S5)**. Because the demographic variables of maternal and paternal age and education, home ownership, and delivery payer are all highly correlated with one another **(Figure 3A)**, it was difficult to identify which traits were the most significant predictors of PCB levels in maternal serum.

To address this question, we computed fitted multiple linear regression models for censored data. First, we identified all PCB-trait correlations with FDR < 0.2. This cut-off was purposely lenient to include all traits that may be important in affecting PCB levels. To address multicollinearity, we removed a PCB-trait pair from analysis if the trait was highly correlated with another trait (r > 0.5) that was more strongly associated (lower FDR) with the PCB congener. With all PCB-trait pairs that met these cut-offs, we performed multiple linear regression using a log scale to normalize the left-censored PCB data. Though all congeners’ multiple linear regression models shared characteristics, each was distinct despite the high correlation between all four PCB congeners/congener pairs **(Figure 2B)**. Maternal age (MomAge) was the most significant predictor (*P* < 0.00001) of maternal levels of PCBs 153 + 168, 170, 180 + 193, and 187, with increasing age predicting increasing serum PCB levels. PCBs 153 + 168 (*P* < 0.05) and 180 + 193 (*P* < 0.005) were also strongly predicted by the child’s birth year (year of maternal serum sample collection), with decreasing levels of PCBs from 2006 to 2016. Higher maternal serum levels of PCB 153 + 168 was also predicted by lower maternal BMI (*P* < 0.05), while higher maternal serum levels of PCB 180 + 193 were predicted by higher levels of maternal plasma polyunsaturated fatty acids (*P* < 0.05) **(Figure 3B)**. Standard errors of the coefficients and other regression model variables are reported in **Supplemental Table S14**.

Fitted multiple linear regression models:

- ln**PCB 153 + 168** = 211 + 0.134\***MomAge** - 0.0322\***MaternalBMI** - 0.109\***BirthYear**^a^
- ln**PCB 170** = -8.9024 + 0.1127\***MomAge**
- ln**PCB 180 + 193** = 219.31014 + 0.09813\***MomAge** + 0.00588\***MaternalPlasmaPUFA** – 0.11286\***BirthYear**^a^
- ln**PCB 187** = -10.1156 + 0.12414\***MomAge**

^a^Birth year refers to child birth year which is also the year of maternal serum sample collection

The multiple linear regression models partially confirmed our hypotheses that child diagnosis of ASD, year of sample collection, and maternal BMI would be significant predictors of maternal serum PCB levels. PCBs 153 + 168, 170, 180 + 193, and 187 were not significantly predicted by ASD diagnosis or ADOS score, which we confirmed by the Peto-Prentice test **(Supplemental Table S15)**. However, the negative associations between PCB levels and year of sample collection as well as maternal BMI confirmed our hypotheses and aligned with previous publications. Due to the negative association between BMI and PCBs, we decided to not lipid-adjust our PCB values. Previous studies have indicated a potential causal effect of PCBs on serum lipid levels (Hennig et al., 2005; Langer et al., 2003), and have found lipid standardization to be prone to bias (Schisterman et al., 2005). Further, different groups of PCBs are differently associated with total serum lipids, making lipid adjustment difficult for all 209 PCB congeners in our study (Aminov et al., 2013).

To investigate why PCB 153 + 168 was more strongly negatively correlated with (and predicted by) maternal BMI than the other three PCB congeners/congener pairs we correlated the BMI-PCB tau value with the octanol-water partition coefficients (K_ow_) for each PCB congener/pair **(Supplemental Figure S6) (Supplemental Table S16)**. K_ow_ represents the lipophilicity of a compound, with more positive values representing increased lipophilicity and decreased water-solubility. Of the four PCB congeners/congeners pairs, PCB 153 + 168 is the least lipophilic (lowest K_ow_) and has the strongest negative PCB-BMI correlation tau value. This inverse relationship between BMI and serum levels of PCBs with lower lipophilicity is revealed among individuals with lower fat stores (lower BMI) that have higher amounts of PCB 153 + 168 in their bloodstream. This may occur with PCB 153 +168 more than the other congeners because its lower lipophilicity, meaning PCB 153 +168 has lower propensity to stay in the fat stores and greater solubility in the bloodstream (Bruno et al., 2021).

Lastly, the multiple linear regression models showed that PCB 180 + 193 serum levels are predicted by maternal plasma polyunsaturated fatty acid levels. PCBs have previously been associated with polyunsaturated fatty acids in human serum due to the high prevalence of both PCBs and PUFAs in fish and fish oil (Ashley et al., 2013; Bourdon et al., 2010; Dellinger et al., 2018; Ge et al., 2019; Tøttenborg et al., 2015). However, we did not include dietary information in this study, so cannot validate a hypothesis for the association between PCB 180 + 193 and dietary polyunsaturated fatty acid intake.

Based on our findings with participant traits and PCBs 153 + 168, 170, 180 + 193, and 187, we assessed detection frequencies **(Figure 3C)** and levels **(Supplemental Figure S7)** of all PCB congeners by offspring sex, offspring ASD diagnosis, maternal age, maternal BMI pre-pregnancy, and offspring birth year (year of maternal serum sample collection) **(Supplemental Table S17)**. Overall, 18 congeners were more frequently detected in women who gave birth at over age 35 (and therefore had their serum collected at over age 35) than those who gave birth at age 35 or younger. One congener was more frequently detected in women with pre-pregnancy BMI 25 or under (PCB 169), while one congener was more frequently detected in women with TD offspring compared to ASD/non-TD (PCB 194). Seven congeners were more frequently detected in those with sample collection (child birth year) between 2006-2011 compared to 2012-2016.

### 3.5. Placental DNA co-methylation modules map to large blocks of adjacent regions

We next assessed DNA methylation patterns from placental samples in the MARBLES cohort to analyze the relationships between DNA methylation, maternal serum PCB levels, and participant traits including those related to child neurodevelopment. Although we did not see a significant association between maternal serum PCB levels and child ASD diagnosis at the population level, placental DNA methylation may act as a measurable intermediary at the level of the individual. To identify patterns of DNA methylation, we used the Comethyl R package (Mordaunt et al., 2022), which performs weighted gene correlation network analysis (WGCNA) with WGBS data. This method allowed us to look at modules of co-methylated regions instead of individual CpG sites, which are more susceptible to stochastic variation. Additionally, Comethyl module eigennodes can be correlated with any number of traits of interest, using a statistical method of choice.

CpGs from 157 placental samples were grouped into regions from which modules were constructed. Correlations between region methylation were all raised to the power of 15 to amplify strong connections and decrease the background noise of weak connections **(Figure 4A)**. Regions formed 6 blocks **(Supplemental Figure S15)** from which 27 modules were identified, plus a grey module, to which regions not assigned to another module were assigned. From 199,945 total filtered regions, 928 were assigned to a module (0.46%) while the remaining regions (99.54%) were assigned to the grey module. Each of the 27 modules contained between 10 and 89 regions **(Figure 4B)**, which is consistent with previous Comethyl findings using cord blood samples (Mordaunt et al., 2022). However, the modules generated from placenta were unique because most modules mapped to large blocks of adjacent regions **(Figure 4C) (Supplemental Table S23)**. This is in contrast to other tissues, where modules typically map to short regions across several chromosomes (Mordaunt et al., 2022). This unique finding led us to investigate where the modules mapped on the genome.

**Figure 4.**
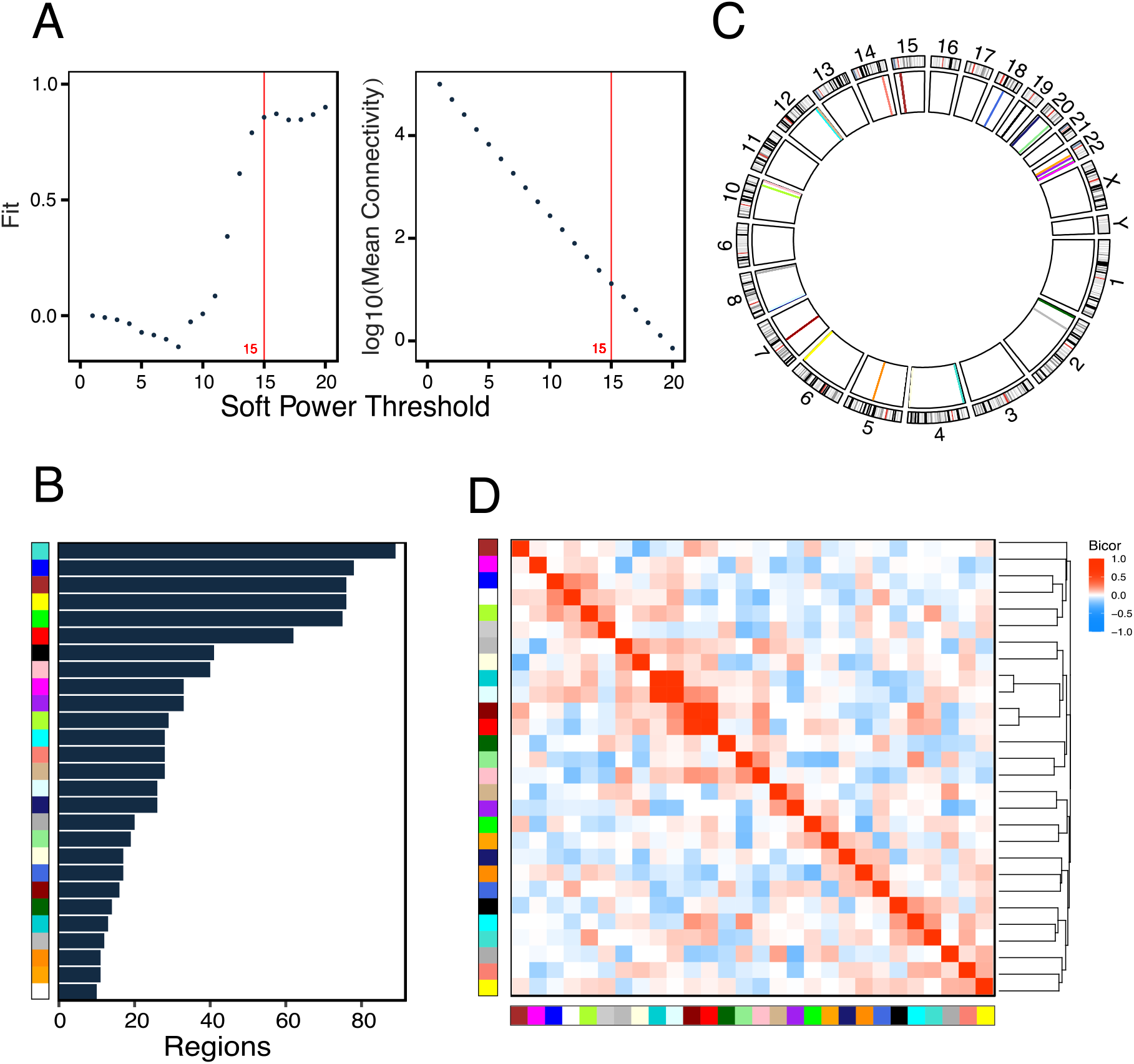
Comethylation network construction and module characterization. **A)** Scale-free topology fit and mean connectivity is plotted at various soft power thresholds. A soft power threshold of 15 was chosen for network construction, meaning all correlations between regions were raised to a power of 15, strengthening strong relationships and reducing background noise caused by weak relationships. **B)** The number of regions assigned to each module ranged from 10 to 89. **C)** Circos plot of regions colored by module in the inner ring and chromosome bands in the outer ring. Most modules map to only one location on the genome due to large blocks of adjacent regions forming modules. **D)** Eigennode values clustered and compared across modules by Bicor. The DarkTurquoise and LightCyan modules are highly correlated, as are DarkRed and Red modules.

### 3.6. Placental DNA co-methylation modules annotate to gene introns and CpG open sea contexts

We tested the hypothesis that co-methylated modules mapped to partially methylated domains (PMDs), regions of the placental genome with reduced average methylation levels (approximately 40-45% methylated) that are highly variable across individuals (Schroeder et al., 2013). Because we selected for regions with a high standard deviation in methylation levels and found that filtered regions with the highest standard deviations had 40-60% mean methylation **(Supplemental Figure S13)**, we hypothesized that these regions would preferentially map to PMDs (Schroeder et al., 2013). Of the 928 regions assigned to modules (excluding the grey module), 407 (43.86%) mapped to PMDs **(Supplemental Table S23)**. PMDs cover an average of 37% of the placental genome (Schroeder et al., 2013), showing that PMDs are over-represented in modules compared to the placental genome (chi-squared *P* < 0.0001) **(Supplemental Table S24)**. Of the 27 modules, 11 consist of 100% regions that map onto PMDs; 9 consist of 100% regions that fall outside of PMDs; and 7 consist of a mixture. The average mean methylation across the regions that do not map to PMDs is 0.6628 (SD 0.1142), while the average mean methylation across the regions that do map to PMDs is 0.6262 (SD 0.1124). These relatively high standard deviations indicate the high variability in methylation levels across regions in both PMDs and non-PMDs.

We then annotated regions within modules to genes, and identified regions’ genic features (i.e. promoter, exon, intron, UTR, enhancer, intergenic) and CpG features (i.e. island, shore, shelf, open sea) **(Supplemental Table S23)**. We found that 877/928 (94.5%) of regions within modules mapped to open sea CpG contexts, supporting previous findings that most DMRs do not reside at CpG islands (Visone et al., 2019). 700/928 (75.4%) of regions in modules mapped to introns of genes, while only 116,878/199,944 (58.5%) of total filtered regions mapped to introns. This indicates that introns are over-represented amongst regions that co-methylate in placenta, compared to background regions (chi-squared *P* < 0.0001) **(Supplemental Table S24)**. The enrichment for introns is likely related to the finding that the entire gene body, including introns and exons, is more highly methylated over active genes in placenta (Schroeder et al, 2016), and introns cover a larger genomic territory than exons.

We also correlated module eigennodes with one another to identify highly similar modules and explore whether they annotated to the same or different genes **(Figure 4D) (Supplemental Figure S16)**. The most highly correlated pairs of modules were DarkTurquoise with LightCyan (Bicor = 0.792, *p*-value = 6.08E-33) and Red with DarkRed (Bicor = 0.718, *p*-value = 1.44E-24). Though these modules are strongly correlated, they were not combined into a single module because they did not meet the Bicor cut-off of 0.9. The DarkTurquoise and LightCyan modules consist of large blocks of adjacent regions that map to *CSMD1*, a gene that codes for a complement control protein. Nearly all of *CSMD1* lies within a placental PMD (Schroeder et al., 2013). Regions in both modules map primarily to *CSMD1* introns with one region from each module mapping to one of *CSDM1*’s 69 exons. The Red and DarkRed modules both consist of large blocks of adjacent regions that map to *AUTS2*, activator of transcription and developmental regulator. The DarkRed and Red regions lie approximately 396-526kb and 364kb-1Mb upstream of *AUTS2*, respectively, showing that regions from the two modules overlap with one another on the genome. Overall, these results highlight the complexity of DNA co-methylation in the placental genome.

### 3.7. Placental DNA co-methylation modules are correlated with child neurodevelopment and map to genes previously associated with ASD, including *GALC, AUTS2*, and *CSMD1*

To explore potential relationships between placental DNA methylation and child neurodevelopment, we correlated module eigennodes with participant traits using Bicor **(Figure 5A)**. We also mapped regions within modules to genes in order to relate methylation in certain genes to traits of interest **(Figure 5C)**. Four modules significantly (*P* < 0.05) correlated with a child neurodevelopmental trait (ASD diagnosis, Mullen Score, or ADOS comparison score): Salmon, DarkRed, Red, and DarkTurquoise **(Figure 6 and Supplemental Figures S17-S19)**. These modules mapped, respectively, to *GALC, AUTS2, AUTS2*, and *CSMD1*, all of which have previously been associated with ASD (Cukier et al., 2014; Hori et al., 2021, p. 2; Zhu et al., 2022).

**Figure 5.**
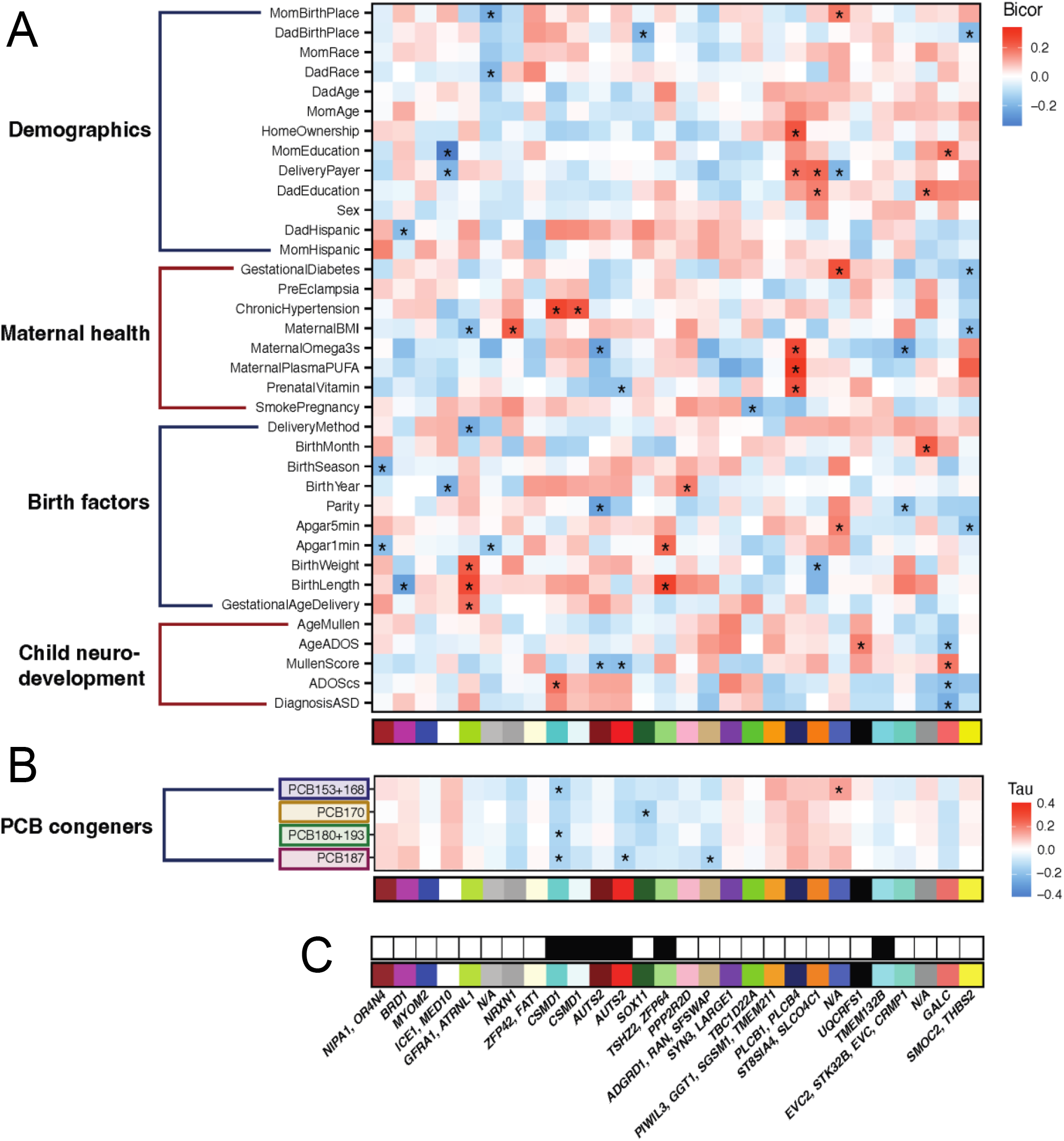
Module eigennodes correlated with **A)** participant traits using biweight midcorrelation (* unadjusted P < 0.05) and **B)** maternal serum PCB levels using Kendall’s tau for left-censored data (* unadjusted P < 0.05). **C)** Modules annotated to genes using the hg38 genome. Black boxes indicate genes that were identified as fetal brain and placenta differentially methylated regions in both sexes in a mouse study of gestational PCB exposure (Laufer et al., 2022)

**Figure 6.**
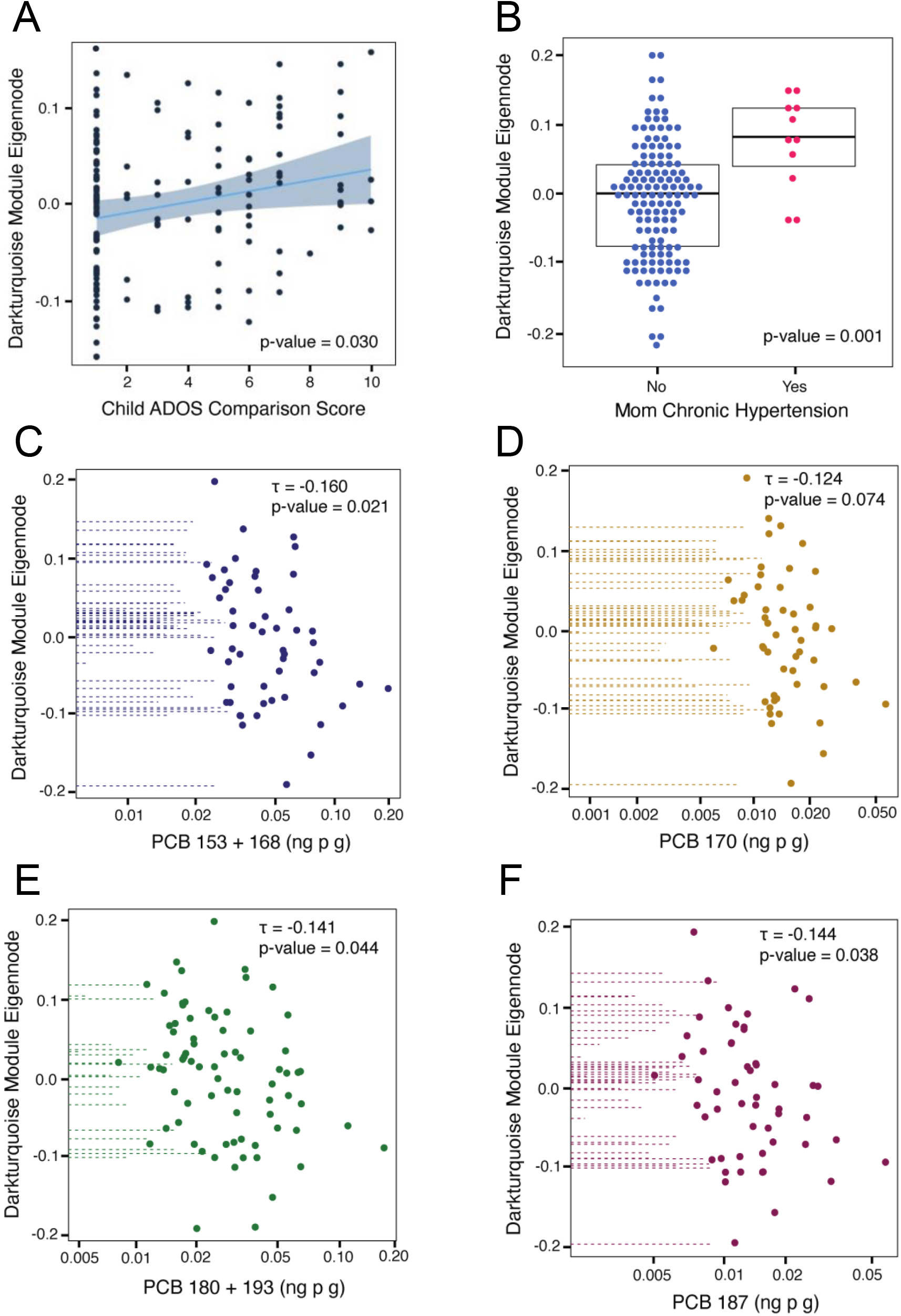
*CSMD1* DarkTurquoise module eigennodes correlated with **A)** Child ADOS comparison score, **B)** mom chronic hypertension and maternal serum levels of PCB **C)** 153 + 168 **D)** 170 **E)** 180 + 193 **F)** 187

The *GALC* Salmon module negatively correlated with ASD diagnosis (*P* < 0.01), as well as ADOS comparison score (*P* < 0.05), but positively correlated with Mullen Score (*P* < 0.05). The Salmon module also positively correlated with maternal education (*P* < 0.05), indicating that increased placental DNA methylation at Salmon loci (*GALC* gene) correlates with more typical development (and less ASD diagnosis) in the offspring and higher maternal education levels. This also fits with the positive correlation of maternal education with the Mullen Score, for which higher values reflect improved cognition and lower ASD diagnosis **(Figure 5A)**. The Salmon module regions map to *GALC*, a gene that encodes the enzyme galactosylceramidase, a component of myelin, the protective coating around some nerve cells. These regions were previously identified as placenta ASD hypomethylated DMRs in the MARBLES cohort using a DMR bioinformatic approach (Zhu et al., 2022).

The *AUTS2* Red and DarkRed modules correlated negatively with Mullen Score, indicating higher methylation at *AUTS2*, a known ASD risk gene, when Mullen Scores are lower and ASD is more likely to be diagnosed. This study may be underpowered to detect a significant correlation between these modules and ASD diagnosis (Red *P* = 0.219, DarkRed *P* = 0.243), a categorical variable (ASD, non-TD, TD), despite their significance with Mullen Score, a continuous variable. Additionally, the correlation with Mullen Score but not with ADOS Score may reflect that methylation at this loci is associated with cognitive measures, as reflected by Mullen, but not with social aspects of ASD, as reflected by ADOS. The *AUTS2* DarkRed module also correlated negatively with maternal omega 3s, while the Red module correlated negatively with prenatal vitamin use, suggesting that high omega 3s and taking prenatal vitamins are associated with lower DNA methylation at this locus, in the expected protective direction for cognition.

The *CSMD1* DarkTurquoise module negatively associated with both ADOS comparison score (*P* < 0.05) and maternal chronic hypertension (*P* < 0.005) **(Figure 6A-B)**. Again, a significant correlation was not detected with ASD diagnosis (*P* = 0.080), potentially because it is a categorical variable, or with Mullen Score, perhaps reflecting the sensitivity of *CSMD1* methylation to the social aspects of ASD that are measured by ADOS rather than the cognitive aspects that are measured by Mullen. Hypertensive disorders during pregnancy have been shown to potentially increase risk of ASD in offspring (Curran et al., 2018) and common variants in *CSMD1* have previously been associated with both hypertension (Hong et al., 2010) and ASD (Cukier et al., 2014), as well as schizophrenia (Liu et al., 2017; Ripke et al., 2014). *CSMD1* has high expression in brain, particularly in neurons (Baum et al., 2020), and was identified as a hypermethylated differentially methylated gene in brain tissue from individuals with Dup15q (Dunaway et al., 2016), a common CNV observed in ASD (Chaste et al., 2014; Hogart et al., 2010).

### 3.8. Placental DNA co-methylation modules are correlated with maternal serum PCBs

To investigate relationships between maternal serum PCBs and placenta DNA methylation, we correlated module eigennodes with PCBs 153 + 168, 170, 180 + 193, and 187 using Kendall’s tau for left-censored data **(Figure 5B)**. The *CSMD1* DarkTurquoise module negatively correlated (*P* < 0.05) with PCBs 153+168, 180+193, and 187 **(Figure 6C-F)**; the *AUTS2* Red (*P* < 0.05) and *ADGRD1* Tan (*P* < 0.05) modules significantly negatively correlated with PCB 187; the *SOX11* DarkGreen module significantly negatively correlated with PCB 170 (*P* < 0.05); and the RoyalBlue module significantly positively correlated with PCB 153 + 168 (*P* < 0.05). Of the seven significant module – PCB correlations, six were negative correlations, meaning that individuals with higher serum PCB levels had lower methylation levels at those loci. These findings align with a mouse model of gestational PCB exposure, in which placenta displayed global hypomethylation in both sexes as well as a large skew in DMRs, where 81% of female and 85% of male DMRs were hypomethylated in PCB-exposed mice compared to controls (Laufer et al., 2022).

Interestingly, the *CSMD1* DarkTurquoise and *AUTS2* Red modules correlated with maternal PCBs as well as measurements of child neurodevelopment. In both modules, the methylation change associated with elevated maternal serum PCB levels appears to be in the opposite direction as ASD in the child, which is the inverse of our hypothesis but accompanies our finding that ASD diagnosis was non-significantly negatively correlated with PCBs 153+168, 170, 180+193, and 187 **(Figure 3B)**. A previous study using a mouse model of prenatal PCB exposure identified DMRs that mapped to *CSMD1* as well as *AUTS2* in male and female placentas and fetal brains **(Figure 5C)** (Laufer et al., 2022). Several DMRs mapped to each gene for both sexes and tissues and were inconsistent in the direction of differential methylation, indicating that PCB exposure may dysregulate DNA methylation at *CSMD1* and *AUTS2* in variable ways. Additionally, methylation levels of *CSMD1* in sperm have been inversely correlated with serum DDE levels in Faroese individuals who are exposed to high levels of organic pollutants (Maggio et al., 2021). DDE and PCB exposures often occur together, and both chemicals contain phenyl groups and chlorine molecules, indicating a potential convergent mechanism.

Additionally, it is notable that the *CSMD1* DarkTurquoise module correlated with maternal chronic hypertension and serum PCB levels **(Figure 6 B-F)**. Hypertension has previously been associated with PCBs, though correlations in the positive (Raffetti et al., 2020) and the negative (Warembourg et al., 2019) direction have both been found. Increases in systolic and diastolic blood pressure have also been previously associated with high fish intake during pregnancy (Warembourg et al., 2019), which is known to increase exposure to PCBs.

### 3.9. Placental DNA co-methylation modules are highly preserved in an independent dataset

Lastly, we assessed the placental DNA methylation module quality and preservation in an independent WGBS ASD and TD placenta dataset from the EARLI study (Langfelder et al., 2011). EARLI networks were constructed using the same set of CpGs and regions as were used in the MARBLES dataset. The four types of features assessed were quality, preservation, accuracy, and separability.

All 27 modules had strong evidence for quality and preservation as shown by all modules having summary.qual and summary.pres Z-scores > 10. 19 modules had strong evidence for accuracy, including two of our modules of interest (*CSMD1* DarkTurquoise and *GALC* Salmon). In contrast, only eight modules showed evidence of separability in the reference set (MARBLES) and one module in the test set (EARLI), which did not include our modules of interest. These results suggest that the identified modules are of high quality and are consistent between multiple placenta datasets **(Supplemental Figure S20)(Supplemental Table S25)**.

## 4. CONCLUSIONS

We measured all PCB congeners in maternal serum collected at delivery from an ASD enriched risk cohort and detected 4 congeners/congener pairs (PCBs 153 + 168, 170, 180 + 193, 187) in more than 50% of samples. While we found maternal age, year of sample collection, pre-pregnancy BMI, and polyunsaturated fatty acid levels to be predictive of maternal serum PCB levels, we did not find an association between maternal PCBs and child ASD diagnosis. This finding may be explained by the fact that PCB congener profiles in pregnant people are changing over time and congeners that affect neuroevelopment may be less prevalent in our population than those in previous studies. Additionally, PCBs may not alter neurodevelopment themselves, but instead modify genetic risk through epigenetic marks such as DNA methylation.

To investigate this potential gene by environment interaction, we used a systems approach to identify regions with correlated DNA methylation in placenta. We identified and replicated 27 co-methylated modules, 4 of which correlated with child neurodevelopment measurements (ASD diagnosis, ADOS comparison score, or Mullen score), and 5 of which correlated with PCB congener levels, as well as other variables, including maternal hypertension and prenatal vitamin use. DNA co-methylation patterns in the placenta identified 2 gene loci (*CSMD1* and *AUTS2*) that were correlated with one or more PCB congeners and child neurodevelopment measures. Interestingly, increased methylation at *CSMD1* and *AUTS2* correlated with higher rates of ASD diagnosis but lower maternal serum PCB levels, indicating complex dysregulation at these loci during development and the need for further investigation to clarify exact mechanisms. Together, these results demonstrate the utility of comparing placental methylation patterns with multiple variables to understand complex gene by environmental interactions in the etiology of ASD and other neurodevelopmental disorders.

## Data Availability

Datasets supporting the conclusions are available in the Gene Expression Omnibus repository (GEO) at accession number (GSE178206). Code and scripts for this study are available on GitHub.

https://github.com/juliamouat/MARBLES_PCB

## ABBREVIATIONS

PCB: polychlorinated biphenyl
WGBS: whole genome bisulfite sequencing
ASD: autism spectrum disorder
TD: typical development
DMR: differentially methylated region
PMD: partially methylated domain
WGCNA: weighted gene correlation network analysis

## USE OF HUMAN SUBJECTS DECLARATION

This study was performed in accordance with the ethical principles for medical research involving human subjects in the Declaration of Helsinki. The University of California, Davis Institutional Review Board and the State of California Committee for the Protection of Human Subjects approved this study and the MARBLES Study protocols (IRB# 225645). Human Subjects Institutional Review Boards at each of the four sites in the EARLI Study approved this study and the EARLI Study protocols (IRB# 214753). Neither data nor specimens were collected until written informed consent to participate was obtained from the parents.

## AUTHOR CONTRIBUTIONS

**Julia S. Mouat:** methodology, software, formal analysis, writing – original draft, visualization, conceptualization **Xueshu Li:** investigation, resources, visualization, writing – review and editing **Kari Neier:** methodology, software, formal analysis, writing – review and editing **Yihui Zhu:** resources, writing – review and editing **Charles E. Mordaunt:** software, writing – review and editing **Michele A. La Merrill:** methodology, writing – review and editing **Hans-Joachim Lehmler:** investigation, resources, supervision, writing – review and editing **Michael P. Jones:** conceptualization, writing – review and editing **Pamela J. Lein:** funding acquisition, supervision, writing – review and editing **Rebecca J. Schmidt:** funding acquisition, supervision, data curation, writing – review and editing **Janine M. LaSalle:** funding acquisition, supervision, conceptualization, writing – review and editing, project administration

## ACKNOWLEDGEMENTS

We would like to thank Anthony E. Valenzuela, University of California, Davis, for joining discussions about this paper and providing helpful background reading; Elizabeth Angel, University of California, Davis, for quickly organizing MARBLES and EARLI metadata; Drs. Rachel F. Marek and Keri C. Hornbuckle, University of Iowa, for supporting the congener specific PCB analysis. The graphical abstract, Figure 1, and Figure 2B were made with BioRender.com.

## REFERENCES

Ahmadi, H., Granger, D. A., Hamilton, K. R., Blair, C., & Riis, J. L. (2021). Censored data considerations and analytical approaches for salivary bioscience data. Psychoneuroendocrinology, 129, 105274. https://doi.org/10.1016/j.psyneuen.2021.105274

Aminov, Z., Haase, R. F., Pavuk, M., & Carpenter, D. O. (2013). Analysis of the effects of exposure to polychlorinated biphenyls and chlorinated pesticides on serum lipid levels in residents of Anniston, Alabama. Environmental Health, 12, 108. https://doi.org/10.1186/1476-069X-12-108

Ashley, J. T. F., Ward, J. S., Anderson, C. S., Schafer, M. W., Zaoudeh, L., Horwitz, R. J., & Velinsky, D. J. (2013). Children’s daily exposure to polychlorinated biphenyls from dietary supplements containing fish oils. Food Additives & Contaminants. Part A, Chemistry, Analysis, Control, Exposure & Risk Assessment, 30(3), 506–514. https://doi.org/10.1080/19440049.2012.753161

Baum, M. L., Wilton, D. K., Muthukumar, A., Fox, R. G., Carey, A., Crotty, W., Scott-Hewitt, N., Bien, E., Sabatini, D. A., Lanser, T., Frouin, A., Gergits, F., Håvik, B., Gialeli, C., Nacu, E., Blom, A. M., Eggan, K., Johnson, M. B., McCarroll, S. A., & Stevens, B. (2020). CUB and Sushi Multiple Domains 1 (CSMD1) opposes the complement cascade in neural tissues (p. 2020.09.11.291427). bioRxiv. https://doi.org/10.1101/2020.09.11.291427

Benjamini, Y., & Hochberg, Y. (1995). Controlling the False Discovery Rate: A Practical and Powerful Approach to Multiple Testing. Journal of the Royal Statistical Society: Series B (Methodological), 57(1), 289–300. https://doi.org/10.1111/j.2517-6161.1995.tb02031.x

Benjamini, Y., & Hochberg, Y. (2000). On the Adaptive Control of the False Discovery Rate in Multiple Testing With Independent Statistics. Journal of Educational and Behavioral Statistics, 25(1), 60–83. https://doi.org/10.3102/10769986025001060

Berghuis, S. A., Soechitram, S. D., Hitzert, M. M., Sauer, P. J. J., & Bos, A. F. (2013). Prenatal exposure to polychlorinated biphenyls and their hydroxylated metabolites is associated with motor development of three-month-old infants. Neurotoxicology, 38, 124–130. https://doi.org/10.1016/j.neuro.2013.07.003

Bernardo, B. A., Lanphear, B. P., Venners, S. A., Arbuckle, T. E., Braun, J. M., Muckle, G., Fraser, W. D., & McCandless, L. C. (2019). Assessing the Relation between Plasma PCB Concentrations and Elevated Autistic Behaviours using Bayesian Predictive Odds Ratios. International Journal of Environmental Research and Public Health, 16(3), 457. https://doi.org/10.3390/ijerph16030457

Bourdon, J. A., Bazinet, T. M., Arnason, T. T., Kimpe, L. E., Blais, J. M., & White, P. A. (2010). Polychlorinated biphenyls (PCBs) contamination and aryl hydrocarbon receptor (AhR) agonist activity of Omega-3 polyunsaturated fatty acid supplements: Implications for daily intake of dioxins and PCBs. Food and Chemical Toxicology: An International Journal Published for the British Industrial Biological Research Association, 48(11), 3093–3097. https://doi.org/10.1016/j.fct.2010.07.051

Brändli, R. C., Bucheli, T. D., Kupper, T., Mayer, J., Stadelmann, F. X., & Tarradellas, J. (2007). Fate of PCBs, PAHs and their source characteristic ratios during composting and digestion of source-separated organic waste in full-scale plants. Environmental Pollution, 148(2), 520–528. https://doi.org/10.1016/j.envpol.2006.11.021

Bruno, C. D., Harmatz, J. S., Duan, S. X., Zhang, Q., Chow, C. R., & Greenblatt, D. J. (2021). Effect of lipophilicity on drug distribution and elimination: Influence of obesity. British Journal of Clinical Pharmacology, 87(8), 3197–3205. https://doi.org/10.1111/bcp.14735

Canales, R. A., Wilson, A. M., Pearce-Walker, J. I., Verhougstraete, M. P., & Reynolds, K. A. (2018). Methods for Handling Left-Censored Data in Quantitative Microbial Risk Assessment. Applied and Environmental Microbiology, 84(20), e01203–18. https://doi.org/10.1128/AEM.01203-18

Cavalcante, R. G., & Sartor, M. A. (2017). annotatr: Genomic regions in context. Bioinformatics, 33(15), 2381–2383. https://doi.org/10.1093/bioinformatics/btx183

CDC. (2022, March 2). Data and Statistics on Autism Spectrum Disorder | CDC. Centers for Disease Control and Prevention. https://www.cdc.gov/ncbddd/autism/data.html

Chaste, P., & Leboyer, M. (2012). Autism risk factors: Genes, environment, and geneenvironment interactions. Dialogues in Clinical Neuroscience, 14(3), 281–292.

Chaste, P., Sanders, S. J., Mohan, K. N., Klei, L., Song, Y., Murtha, M. T., Hus, V., Lowe, J. K., Willsey, A. J., Moreno-De-Luca, D., Yu, T. W., Fombonne, E., Geschwind, D., Grice, D. E., Ledbetter, D. H., Lord, C., Mane, S. M., Martin, D. M., Morrow, E. M., … Kim, S.-J. (2014). Modest impact on risk for autism spectrum disorder of rare copy number variants at 15q11.2, specifically breakpoints 1 to 2. Autism Research : Official Journal of the International Society for Autism Research, 7(3), 355–362. https://doi.org/10.1002/aur.1378

Chawarska, K., Shic, F., Macari, S., Campbell, D. J., Brian, J., Landa, R., Hutman, T., Nelson, C. A., Ozonoff, S., Tager-Flusberg, H., Young, G. S., Zwaigenbaum, L., Cohen, I. L., Charman, T., Messinger, D. S., Klin, A., Johnson, S., & Bryson, S. (2014). 18-month predictors of later outcomes in younger siblings of children with autism spectrum disorder: A baby siblings research consortium study. Journal of the American Academy of Child and Adolescent Psychiatry, 53(12), 1317-1327.e1. https://doi.org/10.1016/j.jaac.2014.09.015

Chen, J. J., Roberson, P. K., & Schell, M. J. (2010). The false discovery rate: A key concept in large-scale genetic studies. Cancer Control: Journal of the Moffitt Cancer Center, 17(1), 58–62. https://doi.org/10.1177/107327481001700108

Cheroni, C., Caporale, N., & Testa, G. (2020). Autism spectrum disorder at the crossroad between genes and environment: Contributions, convergences, and interactions in ASD developmental pathophysiology. Molecular Autism, 11, 69. https://doi.org/10.1186/s13229-020-00370-1

Chu, S., Covaci, A., & Schepens, P. (2003). Levels and chiral signatures of persistent organochlorine pollutants in human tissues from Belgium. Environmental Research, 93(2), 167–176. https://doi.org/10.1016/s0013-9351(03)00016-1

Coletta, J. M., Bell, S. J., & Roman, A. S. (2010). Omega-3 Fatty Acids and Pregnancy. Reviews in Obstetrics and Gynecology, 3(4), 163–171.

Corrigan, F. M., Murray, L., Wyatt, C. L., & Shore, R. F. (1998). Diorthosubstituted polychlorinated biphenyls in caudate nucleus in Parkinson’s disease. Experimental Neurology, 150(2), 339–342. https://doi.org/10.1006/exnr.1998.6776

Cukier, H. N., Dueker, N. D., Slifer, S. H., Lee, J. M., Whitehead, P. L., Lalanne, E., Leyva, N., Konidari, I., Gentry, R. C., Hulme, W. F., Booven, D. V., Mayo, V., Hofmann, N. K., Schmidt, M. A., Martin, E. R., Haines, J. L., Cuccaro, M. L., Gilbert, J. R., & Pericak-Vance, M. A. (2014). Exome sequencing of extended families with autism reveals genes shared across neurodevelopmental and neuropsychiatric disorders. Molecular Autism, 5, 1. https://doi.org/10.1186/2040-2392-5-1

Curran, E. A., O’Keeffe, G. W., Looney, A. M., Moloney, G., Hegarty, S. V., Murray, D. M., Khashan, A. S., & Kenny, L. C. (2018). Exposure to Hypertensive Disorders of Pregnancy Increases the Risk of Autism Spectrum Disorder in Affected Offspring. Molecular Neurobiology, 55(7), 5557–5564. https://doi.org/10.1007/s12035-017-0794-x

Dellinger, M. J., Olson, J. T., Holub, B. J., & Ripley, M. P. (2018). MERCURY, POLYCHORLINATED BIPHENYLS, SELENIUM, AND FATTY ACIDS IN TRIBAL FISH HARVESTS OF THE UPPER GREAT LAKES. Risk Analysis : An Official Publication of the Society for Risk Analysis, 38(10), 2029–2040. https://doi.org/10.1111/risa.13112

DeVoto, E., Fiore, B. J., Millikan, R., Anderson, H. A., Sheldon, L., Sonzogni, W. C., & Longnecker, M. P. (1997). Correlations among human blood levels of specific PCB congeners and implications for epidemiologic studies. American Journal of Industrial Medicine, 32(6), 606–613. https://doi.org/10.1002/(sici)1097-0274(199712)32:6<606::aid-ajim6>3.0.co;2-n

Dewailly, E., Mulvad, G., Pedersen, H. S., Ayotte, P., Demers, A., Weber, J. P., & Hansen, J. C. (1999). Concentration of organochlorines in human brain, liver, and adipose tissue autopsy samples from Greenland. Environmental Health Perspectives, 107(10), 823–828. https://doi.org/10.1289/ehp.99107823

Domingo, J. L., & Bocio, A. (2007). Levels of PCDD/PCDFs and PCBs in edible marine species and human intake: A literature review. Environment International, 33(3), 397–405. https://doi.org/10.1016/j.envint.2006.12.004

Dong, H.-Y., Feng, J.-Y., Li, H.-H., Yue, X.-J., & Jia, F.-Y. (2022). Non-parental caregivers, low maternal education, gastrointestinal problems and high blood lead level: Predictors related to the severity of autism spectrum disorder in Northeast China. BMC Pediatrics, 22(1), 11. https://doi.org/10.1186/s12887-021-03086-0

Dunaway, K. W., Islam, M. S., Coulson, R. L., Lopez, S. J., Vogel Ciernia, A., Chu, R. G., Yasui, D. H., Pessah, I. N., Lott, P., Mordaunt, C., Meguro-Horike, M., Horike, S.-I., Korf, I., & LaSalle, J. M. (2016). Cumulative Impact of Polychlorinated Biphenyl and Large Chromosomal Duplications on DNA Methylation, Chromatin, and Expression of Autism Candidate Genes. Cell Reports, 17(11), 3035–3048. https://doi.org/10.1016/j.celrep.2016.11.058

Gabryszewska, M., & Gworek, B. (2021). Municipal waste landfill as a source of polychlorinated biphenyls releases to the environment. PeerJ, 9, e10546. https://doi.org/10.7717/peerj.10546

Ge, F., Wang, X., Zhang, K., Jin, X., Guo, R., Liu, Y., Qiao, X., Zhao, X., Zheng, B., & Zheng, X. (2019). The correlation study between PCBs and δ15N values or FAs in fish collected from Dongting Lake. Chemosphere, 234, 763–768. https://doi.org/10.1016/j.chemosphere.2019.06.094

Granillo, L., Sethi, S., Keil, K. P., Lin, Y., Ozonoff, S., Iosif, A.-M., Puschner, B., & Schmidt, R. J. (2019). Polychlorinated biphenyls influence on autism spectrum disorder risk in the MARBLES cohort. Environmental Research, 171, 177–184. https://doi.org/10.1016/j.envres.2018.12.061

Grimm, F., Hu, D., Kania-Korwel, I., Lehmler, H., Ludewig, G., Hornbuckle, K., Duffel, M., Bergman, A., & Robertson, L. (2015). Metabolism and metabolites of polychlorinated biphenyls (PCBs). Critical Reviews in Toxicology, 45(3), 245–272. https://doi.org/10.3109/10408444.2014.999365

Gupta, P., Thompson, B. L., Wahlang, B., Jordan, C. T., Hilt, J. Z., Hennig, B., & Dziubla, T. (2018). The Environmental Pollutant, Polychlorinated Biphenyls, and Cardiovascular Disease: A Potential Target for Antioxidant Nanotherapeutics. Drug Delivery and Translational Research, 8(3), 740–759. https://doi.org/10.1007/s13346-017-0429-9

Guvenius, D. M., Aronsson, A., Ekman-Ordeberg, G., Bergman, A., & Norén, K. (2003). Human prenatal and postnatal exposure to polybrominated diphenyl ethers, polychlorinated biphenyls, polychlorobiphenylols, and pentachlorophenol. Environmental Health Perspectives, 111(9), 1235–1241. https://doi.org/10.1289/ehp.5946

Hawker, D. W., & Connell, D. W. (1988). Octanol-water partition coefficients of polychlorinated biphenyl congeners. Environmental Science & Technology, 22(4), 382–387. https://doi.org/10.1021/es00169a004

Helsel, D. R. (2005). Nondetects and data analysis: Statistics for censored environmental data. John Wiley & Sons.

Hennig, B., Reiterer, G., Toborek, M., Matveev, S. V., Daugherty, A., Smart, E., & Robertson, L. W. (2005). Dietary fat interacts with PCBs to induce changes in lipid metabolism in mice deficient in low-density lipoprotein receptor. Environmental Health Perspectives, 113(1), 83–87. https://doi.org/10.1289/ehp.7280

Herrick, R. F., Stewart, J. H., & Allen, J. G. (2016). Review of PCBs in US Schools: A Brief History, Estimate of the Number of Impacted Schools, and an Approach for Evaluating Indoor Air Samples. Environmental Science and Pollution Research International, 23(3), 1975–1985. https://doi.org/10.1007/s11356-015-4574-8

Hertz-Picciotto, I., Schmidt, R. J., Walker, C. K., Bennett, D. H., Oliver, M., Shedd-Wise, K. M., LaSalle, J. M., Giulivi, C., Puschner, B., Thomas, J., Roa, D. L., Pessah, I. N., Van de Water, J., Tancredi, D. J., & Ozonoff, S. (2018). A Prospective Study of Environmental Exposures and Early Biomarkers in Autism Spectrum Disorder: Design, Protocols, and Preliminary Data from the MARBLES Study. Environmental Health Perspectives, 126(11), 117004. https://doi.org/10.1289/EHP535

Hogart, A., Wu, D., LaSalle, J. M., & Schanen, N. C. (2010). The Comorbidity of Autism with the Genomic Disorders of Chromosome 15q11.2-q13. Neurobiology of Disease, 38(2), 181–191. https://doi.org/10.1016/j.nbd.2008.08.011

Hong, K.-W., Go, M. J., Jin, H.-S., Lim, J.-E., Lee, J.-Y., Han, B. G., Hwang, S.-Y., Lee, S.-H., Park, H. K., Cho, Y. S., & Oh, B. (2010). Genetic variations in ATP2B1, CSK, ARSG and CSMD1 loci are related to blood pressure and/or hypertension in two Korean cohorts. Journal of Human Hypertension, 24(6), 367–372. https://doi.org/10.1038/jhh.2009.86

Hori, K., Shimaoka, K., & Hoshino, M. (2021). AUTS2 Gene: Keys to Understanding the Pathogenesis of Neurodevelopmental Disorders. Cells, 11(1), 11. https://doi.org/10.3390/cells11010011

Hu, D., & Hornbuckle, K. C. (2010). Inadvertent Polychlorinated Biphenyls in Commercial Paint Pigments. Environmental Science & Technology, 44(8), 2822–2827. https://doi.org/10.1021/es902413k

Huang, Y., Iosif, A.-M., Hansen, R. L., & Schmidt, R. J. (2020). Maternal polyunsaturated fatty acids and risk for autism spectrum disorder in the MARBLES high-risk study. Autism: The International Journal of Research and Practice, 24(5), 1191–1200. https://doi.org/10.1177/1362361319877792

Jeong, Y., Lee, S., Kim, S., Park, J., Kim, H.-J., Choi, G., Choi, S., Kim, S., Kim, S. Y., Kim, S., Choi, K., & Moon, H.-B. (2018). Placental transfer of persistent organic pollutants and feasibility using the placenta as a non-invasive biomonitoring matrix. The Science of the Total Environment, 612, 1498–1505. https://doi.org/10.1016/j.scitotenv.2017.07.054

Jones, K. C., Sanders, G., Wild, S. R., Burnett, V., & Johnston, A. E. (1992). Evidence for a decline of PCBs and PAHs in rural vegetation and air in the United Kingdom. Nature, 356(6365), Article 6365. https://doi.org/10.1038/356137a0

Kashyap, M. L., Sivasamboo, R., Sothy, S. P., Cheah, J. S., & Gartside, P. S. (1976). Carbohydrate and lipid metabolism during human labor: Free fatty acids, glucose, insulin, and lactic acid metabolism during normal and oxytocin-induced labor for postmaturity. Metabolism: Clinical and Experimental, 25(8), 865–875. https://doi.org/10.1016/0026-0495(76)90119-0

Kato, S., McKinney, J. D., & Matthews, H. B. (1980). Metabolism of symmetrical hexachlorobiphenyl isomers in the rat. Toxicology and Applied Pharmacology, 53(3), 389–398. https://doi.org/10.1016/0041-008x(80)90352-x

Koh, W. X., Hornbuckle, K. C., Wang, K., & Thorne, P. S. (2016). Serum Polychlorinated Biphenyls and Their Hydroxylated Metabolites are Associated with Demographic and Behavioral Factors in Children and Mothers. Environment International, 94, 538–545. https://doi.org/10.1016/j.envint.2016.06.014

Korthauer, K., Kimes, P. K., Duvallet, C., Reyes, A., Subramanian, A., Teng, M., Shukla, C., Alm, E. J., & Hicks, S. C. (2019). A practical guide to methods controlling false discoveries in computational biology. Genome Biology, 20(1), 118. https://doi.org/10.1186/s13059-019-1716-1

Ladd-Acosta, C., Andrews, S. V., Bakulski, K. M., Feinberg, J. I., Tryggvadottir, R., Yao, R., Croen, L. A., Hertz-Picciotto, I., Newschaffer, C. J., Salafia, C. M., Feinberg, A. P., Hansen, K. D., & Daniele Fallin, M. (2021). Placenta DNA methylation at ZNF300 is associated with fetal sex and placental morphology [Preprint]. Genomics. https://doi.org/10.1101/2021.03.05.433992

Lan, T., Liu, B., Bao, W., & Thorne, P. S. (2021). BMI modifies the association between dietary intake and serum levels of PCBs. Environment International, 156, 106626. https://doi.org/10.1016/j.envint.2021.106626

Langer, P., Kocan, A., Tajtáková, M., Petrík, J., Chovancová, J., Drobná, B., Jursa, S., Pavúk, M., Koska, J., Trnovec, T., Seböková, E., & Klimes, I. (2003). Possible effects of polychlorinated biphenyls and organochlorinated pesticides on the thyroid after long-term exposure to heavy environmental pollution. Journal of Occupational and Environmental Medicine, 45(5), 526–532. https://doi.org/10.1097/01.jom.0000058346.05741.b0

Langfelder, P., & Horvath, S. (2008). WGCNA: An R package for weighted correlation network analysis. BMC Bioinformatics, 9(1), 559. https://doi.org/10.1186/1471-2105-9-559

Langfelder, P., Luo, R., Oldham, M. C., & Horvath, S. (2011). Is My Network Module Preserved and Reproducible? PLoS Computational Biology, 7(1), e1001057. https://doi.org/10.1371/journal.pcbi.1001057

Laufer, B. I., Neier, K., Valenzuela, A. E., Yasui, D. H., Schmidt, R. J., Lein, P. J., & LaSalle, J. M. (2022). Placenta and fetal brain share a neurodevelopmental disorder DNA methylation profile in a mouse model of prenatal PCB exposure. Cell Reports, 38(9), 110442. https://doi.org/10.1016/j.celrep.2022.110442

Lee, L. (2020). NADA: Nondetects and Data Analysis for Environmental Data. R package version 1.6-1.1. https://CRAN.R-project.org/package=NADA

Lein, P. J. (2015). Chapter 1—Overview of the Role of Environmental Factors in Neurodevelopmental Disorders. In M. Aschner & L. G. Costa (Eds.), Environmental Factors in Neurodevelopmental and Neurodegenerative Disorders (pp. 3–20). Academic Press. https://doi.org/10.1016/B978-0-12-800228-5.00001-7

Leng, L., Li, J., Luo, X., Kim, J., Li, Y., Guo, X., Chen, X., Yang, Q., Li, G., & Tang, N. (2016). Polychlorinated biphenyls and breast cancer: A congener-specific meta-analysis. Environment International, 88, 133–141. https://doi.org/10.1016/j.envint.2015.12.022

Leonard, H., Petterson, B., De Klerk, N., Zubrick, S. R., Glasson, E., Sanders, R., & Bower, C. (2005). Association of sociodemographic characteristics of children with intellectual disability in Western Australia. Social Science & Medicine, 60(7), 1499–1513. https://doi.org/10.1016/j.socscimed.2004.08.014

Levine, S. Z., Kodesh, A., Viktorin, A., Smith, L., Uher, R., Reichenberg, A., & Sandin, S. (2018). Association of Maternal Use of Folic Acid and Multivitamin Supplements in the Periods Before and During Pregnancy With the Risk of Autism Spectrum Disorder in Offspring. JAMA Psychiatry, 75(2), 176–184. https://doi.org/10.1001/jamapsychiatry.2017.4050

Li, X., Hefti, M. M., Marek, R. F., Hornbuckle, K. C., Wang, K., & Lehmler, H.-J. (2022). Assessment of Polychlorinated Biphenyls and Their Hydroxylated Metabolites in Postmortem Human Brain Samples: Age and Brain Region Differences. Environmental Science & Technology, acs.est.2c00581. https://doi.org/10.1021/acs.est.2c00581

Li, X., Westra, B., Behan-Bush, R. M., Liszewski, J. N., Schrodt, M. V., Vats, B., Klingelhutz, A. J., Ankrum, J. A., & Lehmler, H.-J. (2022). Dataset for synthesis and characterization of Cabinet Mixture—University of Iowa. https://iro.uiowa.edu/esploro/outputs/dataset/9984274149002771

Liu, W., Liu, F., Xu, X., & Bai, Y. (2017). Replicated association between the European GWAS locus rs10503253 at CSMD1 and schizophrenia in Asian population. Neuroscience Letters, 647, 122–128. https://doi.org/10.1016/j.neulet.2017.03.039

Lord, C., Risi, S., Lambrecht, L., Cook, E. H., Leventhal, B. L., DiLavore, P. C., Pickles, A., & Rutter, M. (2000). The autism diagnostic observation schedule-generic: A standard measure of social and communication deficits associated with the spectrum of autism. Journal of Autism and Developmental Disorders, 30(3), 205–223.

Lord, C., Rutter, M., & Le Couteur, A. (1994). Autism Diagnostic Interview-Revised: A revised version of a diagnostic interview for caregivers of individuals with possible pervasive developmental disorders. Journal of Autism and Developmental Disorders, 24(5), 659–685. https://doi.org/10.1007/BF02172145

Louis, C., Covaci, A., Crocker, D. E., & Debier, C. (2016). Lipophilicity of PCBs and fatty acids determines their mobilisation from blubber of weaned northern elephant seal pups. Science of The Total Environment, 541, 599–602. https://doi.org/10.1016/j.scitotenv.2015.09.094

Lyall, K., Croen, L., Daniels, J., Fallin, M. D., Ladd-Acosta, C., Lee, B. K., Park, B. Y., Snyder, N. W., Schendel, D., Volk, H., Windham, G. C., & Newschaffer, C. (2017). The Changing Epidemiology of Autism Spectrum Disorders. Annual Review of Public Health, 38, 81–102. https://doi.org/10.1146/annurev-publhealth-031816-044318

Lyall, K., Croen, Lisa. A., Sjödin, A., Yoshida, C. K., Zerbo, O., Kharrazi, M., & Windham, G. C. (2017). Polychlorinated Biphenyl and Organochlorine Pesticide Concentrations in Maternal Mid-Pregnancy Serum Samples: Association with Autism Spectrum Disorder and Intellectual Disability. Environmental Health Perspectives, 125(3), 474–480. https://doi.org/10.1289/EHP277

Maenner, M. J. (2021). Prevalence and Characteristics of Autism Spectrum Disorder Among Children Aged 8 Years—Autism and Developmental Disabilities Monitoring Network, 11 Sites, United States, 2018. MMWR. Surveillance Summaries, 70. https://doi.org/10.15585/mmwr.ss7011a1

Maggio, A. G., Shu, H. T., Laufer, B. I., Hwang, H., Bi, C., Lai, Y., LaSalle, J. M., & Hu, V. W. (2021). Impact of exposures to persistent endocrine disrupting compounds on the sperm methylome in regions associated with neurodevelopmental disorders (p. 2021.02.21.21252162). medRxiv. https://doi.org/10.1101/2021.02.21.21252162

Marek, R. F., Thorne, P. S., DeWall, J., & Hornbuckle, K. C. (2014). Variability in PCB and OH-PCB Serum Levels in Children and Their Mothers in Urban and Rural U.S. Communities. Environmental Science & Technology, 48(22), 13459–13467. https://doi.org/10.1021/es502490w

Mathews, H. B., & Anderson, M. W. (1975). Effect of chlorination on the distribution and excretion of polychlorinated biphenyls. Drug Metabolism and Disposition: The Biological Fate of Chemicals, 3(5), 371–380.

McLean, C. Y., Bristor, D., Hiller, M., Clarke, S. L., Schaar, B. T., Lowe, C. B., Wenger, A. M., & Bejerano, G. (2010). GREAT improves functional interpretation of cis-regulatory regions. Nature Biotechnology, 28(5), 495–501. https://doi.org/10.1038/nbt.1630

Milanowski, B., Lulek, J., Lehmler, H.-J., & Kania-Korwel, I. (2010). Assessment of the Disposition of Chiral Polychlorinated Biphenyls in Female mdr 1a/b Knockout versus Wild-type Mice Using Multivariate Analyses. Environment International, 36(8), 884–892. https://doi.org/10.1016/j.envint.2009.10.007

Mills, R. A., Millis, C. D., Dannan, G. A., Guengerich, F. P., & Aust, S. D. (1985). Studies on the structure-activity relationships for the metabolism of polybrominated biphenyls by rat liver microsomes. Toxicology and Applied Pharmacology, 78(1), 96–104. https://doi.org/10.1016/0041-008x(85)90309-6

Mitchell, M. M., Woods, R., Chi, L.-H., Schmidt, R. J., Pessah, I. N., Kostyniak, P. J., & LaSalle, J. M. (2012). Levels of select PCB and PBDE congeners in human post-mortem brain reveal possible environmental involvement in 15q11-q13 duplication autism spectrum disorder. Environmental and Molecular Mutagenesis, 53(8), 589–598. https://doi.org/10.1002/em.21722

Moore, L. D., Le, T., & Fan, G. (2013). DNA Methylation and Its Basic Function. Neuropsychopharmacology, 38(1), 23–38. https://doi.org/10.1038/npp.2012.112

Mordaunt, C. E., Jianu, J. M., Laufer, B. I., Zhu, Y., Hwang, H., Dunaway, K. W., Bakulski, K. M., Feinberg, J. I., Volk, H. E., Lyall, K., Croen, L. A., Newschaffer, C. J., Ozonoff, S., Hertz-Picciotto, I., Fallin, M. D., Schmidt, R. J., & LaSalle, J. M. (2020). Cord blood DNA methylome in newborns later diagnosed with autism spectrum disorder reflects early dysregulation of neurodevelopmental and X-linked genes. Genome Medicine, 12. https://doi.org/10.1186/s13073-020-00785-8

Mordaunt, C. E., Mouat, J. S., Schmidt, R. J., & LaSalle, J. M. (2022). Comethyl: A network-based methylome approach to investigate the multivariate nature of health and disease. Briefings in Bioinformatics, bbab554. https://doi.org/10.1093/bib/bbab554

Mori, C., Kakuta, K., Matsuno, Y., Todaka, E., Watanabe, M., Hanazato, M., Kawashiro, Y., & Fukata, H. (2014). Polychlorinated biphenyl levels in the blood of Japanese individuals ranging from infants to over 80 years of age. Environmental Science and Pollution Research International, 21(10), 6434–6439. https://doi.org/10.1007/s11356-013-1965-6

Mullen, E. M. (1995). Mullen scales of early learning. AGS Circle Pines, MN.

Mullerova, D., Kopecky, J., Matejkova, D., Muller, L., Rosmus, J., Racek, J., Sefrna, F., Opatrna, S., Kuda, O., & Matejovic, M. (2008). Negative association between plasma levels of adiponectin and polychlorinated biphenyl 153 in obese women under nonenergy-restrictive regime. International Journal of Obesity (2005), 32(12), 1875–1878. https://doi.org/10.1038/ijo.2008.169

Naqvi, A., Qadir, A., Mahmood, A., Baqar, M., Aslam, I., Sajid, F., Mumtaz, M., Li, J., & Zhang, G. (2018). Quantification of polychlorinated biphenyl contamination using human placenta as biomarker from Punjab Province, Pakistan. Environmental Science and Pollution Research, 25(15), 14551–14562. https://doi.org/10.1007/s11356-018-1535-z

Newschaffer, C. J., Croen, L. A., Fallin, M. D., Hertz-Picciotto, I., Nguyen, D. V., Lee, N. L., Berry, C. A., Farzadegan, H., Hess, H. N., Landa, R. J., Levy, S. E., Massolo, M. L., Meyerer, S. C., Mohammed, S. M., Oliver, M. C., Ozonoff, S., Pandey, J., Schroeder, A., & Shedd-Wise, K. M. (2012). Infant siblings and the investigation of autism risk factors. Journal of Neurodevelopmental Disorders, 4(1), 7. https://doi.org/10.1186/1866-1955-4-7

Ouidir, M., Mendola, P., Buck Louis, G. M., Kannan, K., Zhang, C., & Tekola-Ayele, F. (2020). Concentrations of persistent organic pollutants in maternal plasma and epigenome-wide placental DNA methylation. Clinical Epigenetics, 12, 103. https://doi.org/10.1186/s13148-020-00894-6

Ozonoff, S., Young, G. S., Belding, A., Hill, M., Hill, A., Hutman, T., Johnson, S., Miller, M., Rogers, S. J., Schwichtenberg, A. J., Steinfeld, M., & Iosif, A.-M. (2014). The Broader Autism Phenotype in Infancy: When Does It Emerge? Journal of the American Academy of Child & Adolescent Psychiatry, 53(4), 398-407.e2. https://doi.org/10.1016/j.jaac.2013.12.020

Panesar, H. K., Kennedy, C. L., Keil Stietz, K. P., & Lein, P. J. (2020). Polychlorinated Biphenyls (PCBs): Risk Factors for Autism Spectrum Disorder? Toxics, 8(3). https://doi.org/10.3390/toxics8030070

Park, J.-S., Bergman, Å., Linderholm, L., Athanasiadou, M., Kocan, A., Petrik, J., Drobna, B., Trnovec, T., Charles, M. J., & Hertz-Picciotto, I. (2008). Placental Transfer of Polychlorinated Biphenyls, Their Hydroxylated Metabolites and Pentachlorophenol in Pregnant Women from Eastern Slovakia. Chemosphere, 70(9), 1676–1684. https://doi.org/10.1016/j.chemosphere.2007.07.049

Parsana, P., Ruberman, C., Jaffe, A. E., Schatz, M. C., Battle, A., & Leek, J. T. (2019). Addressing confounding artifacts in reconstruction of gene co-expression networks. Genome Biology, 20(1), 94. https://doi.org/10.1186/s13059-019-1700-9

Pauwels, A., Covaci, A., Delbeke, L., Punjabi, U., & Schepens, P. J. C. (1999). The relation between levels of selected PCB congeners in human serum and follicular fluid. Chemosphere, 39(14), 2433–2441. https://doi.org/10.1016/S0045-6535(99)00170-8

Pessah, I. N., Lein, P. J., Seegal, R. F., & Sagiv, S. K. (2019). Neurotoxicity of Polychlorinated Biphenyls and Related Organohalogens. Acta Neuropathologica, 138(3), 363–387. https://doi.org/10.1007/s00401-019-01978-1

Raffetti, E., Donato, F., De Palma, G., Leonardi, L., Sileo, C., & Magoni, M. (2020). Polychlorinated biphenyls (PCBs) and risk of hypertension: A population-based cohort study in a North Italian highly polluted area. The Science of the Total Environment, 714, 136660. https://doi.org/10.1016/j.scitotenv.2020.136660

Rattan, S., Zhou, C., Chiang, C., Mahalingam, S., Brehm, E., & Flaws, J. A. (2017). Exposure to endocrine disruptors during adulthood: Consequences for female fertility. The Journal of Endocrinology, 233(3), R109–R129. https://doi.org/10.1530/JOE-17-0023

Revelle, W. (2022). psych: Procedures for Psychological, Psychometric, and Personality Research (2.2.5). Northwestern University, Evanston, Illinois. https://CRAN.R-project.org/package=psych

Rhind, S. M. (2002). Endocrine disrupting compounds and farm animals: Their properties, actions and routes of exposure. Domestic Animal Endocrinology, 23(1–2), 179–187. https://doi.org/10.1016/s0739-7240(02)00155-8

Ripke, S., Neale, B. M., Corvin, A., Walters, J. T., Farh, K.-H., Holmans, P. A., Lee, P., Bulik-Sullivan, B., Collier, D. A., Huang, H., Pers, T. H., Agartz, I., Agerbo, E., Albus, M., Alexander, M., Amin, F., Bacanu, S. A., Begemann, M., Belliveau, R. A., … O’Donovan, M. C. (2014). Biological Insights From 108 Schizophrenia-Associated Genetic Loci. Nature, 511(7510), 421–427. https://doi.org/10.1038/nature13595

Saktrakulkla, P., Lan, T., Hua, J., Marek, R. F., Thorne, P. S., & Hornbuckle, K. C. (2020). Polychlorinated Biphenyls in Food. Environmental Science & Technology, 54(18), 11443–11452. https://doi.org/10.1021/acs.est.0c03632

Schisterman, E. F., Whitcomb, B. W., Louis, G. M. B., & Louis, T. A. (2005). Lipid adjustment in the analysis of environmental contaminants and human health risks. Environmental Health Perspectives, 113(7), 853–857. https://doi.org/10.1289/ehp.7640

Schmidt, R. J., Hansen, R. L., Hartiala, J., Allayee, H., Schmidt, L. C., Tancredi, D. J., Tassone, F., & Hertz-Picciotto, I. (2011). Prenatal vitamins, one-carbon metabolism gene variants, and risk for autism. Epidemiology (Cambridge, Mass.), 22(4), 476–485. https://doi.org/10.1097/EDE.0b013e31821d0e30

Schmidt, R. J., Iosif, A.-M., Guerrero Angel, E., & Ozonoff, S. (2019). Association of Maternal Prenatal Vitamin Use With Risk for Autism Spectrum Disorder Recurrence in Young Siblings. JAMA Psychiatry, 76(4), 391–398. https://doi.org/10.1001/jamapsychiatry.2018.3901

Schoultz, P., Landoll, J., Coman, D., Gutierrez, A., Alessandri, M., Hume, K., Sperry, L., Boyd, B., & Odom, S. (2010, May 20). The Role of Maternal Education and Stress On Developmental Rates for Preschool Children with Autism Spectrum Disorders.

Schroeder, D. I., Blair, J. D., Lott, P., Yu, H. O. K., Hong, D., Crary, F., Ashwood, P., Walker, C., Korf, I., Robinson, W. P., & LaSalle, J. M. (2013). The human placenta methylome. Proceedings of the National Academy of Sciences of the United States of America, 110(15), 6037–6042. https://doi.org/10.1073/pnas.1215145110

Sethi, S., Morgan, R. K., Feng, W., Lin, Y., Li, X., Luna, C., Koch, M., Bansal, R., Duffel, M. W., Puschner, B., Zoeller, R. T., Lehmler, H.-J., Pessah, I. N., & Lein, P. J. (2019). Comparative Analyses of the 12 Most Abundant PCB Congeners Detected in Human Maternal Serum for Activity at the Thyroid Hormone Receptor and Ryanodine Receptor. Environmental Science & Technology, 53(7), 3948–3958. https://doi.org/10.1021/acs.est.9b00535

Soechitram, S. D., Athanasiadou, M., Hovander, L., Bergman, Å., & Sauer, P. J. J. (2004). Fetal Exposure to PCBs and Their Hydroxylated Metabolites in a Dutch Cohort. Environmental Health Perspectives, 112(11), 1208–1212. https://doi.org/10.1289/ehp.6424

Stockholm Convention. (2001). PCBs—Overview. http://chm.pops.int/implementation/pcbs/overview/tabid/273/default.aspx

Tøttenborg, S. S., Choi, A. L., Bjerve, K. S., Weihe, P., & Grandjean, P. (2015). Effect of seafood mediated PCB exposure on desaturase activity and PUFA profile in Faroese septuagenarians. Environmental Research, 140, 699–703. https://doi.org/10.1016/j.envres.2015.06.001

US EPA, O. (2015, September 25). Table of Polychlorinated Biphenyl (PCB) Congeners [Overviews and Factsheets]. https://www.epa.gov/pcbs/table-polychlorinated-biphenyl-pcb-congeners

Visone, R., Bacalini, M. G., Franco, S. D., Ferracin, M., Colorito, M. L., Pagotto, S., Laprovitera, N., Licastro, D., Marco, M. D., Scavo, E., Bassi, C., Saccenti, E., Nicotra, A., Grzes, M., Garagnani, P., Laurenzi, V. D., Valeri, N., Mariani-Costantini, R., Negrini, M., … Veronese, A. (2019). DNA methylation of shelf, shore and open sea CpG positions distinguish high microsatellite instability from low or stable microsatellite status colon cancer stem cells. Epigenomics, 11(6), 587–604. https://doi.org/10.2217/epi-2018-0153

Wagner, C. C., Amos, H. M., Thackray, C. P., Zhang, Y., Lundgren, E. W., Forget, G., Friedman, C. L., Selin, N. E., Lohmann, R., & Sunderland, E. M. (2019). A Global 3-D Ocean Model for PCBs: Benchmark Compounds for Understanding the Impacts of Global Change on Neutral Persistent Organic Pollutants. Global Biogeochemical Cycles, 33(3), 469–481. https://doi.org/10.1029/2018GB006018

Wan, M. L. Y., Co, V. A., & El-Nezami, H. (2022). Endocrine disrupting chemicals and breast cancer: A systematic review of epidemiological studies. Critical Reviews in Food Science and Nutrition, 62(24), 6549–6576. https://doi.org/10.1080/10408398.2021.1903382

Warembourg, C., Maitre, L., Tamayo-Uria, I., Fossati, S., Roumeliotaki, T., Aasvang, G. M., Andrusaityte, S., Casas, M., Cequier, E., Chatzi, L., Dedele, A., Gonzalez, J.-R., Gražulevičienė, R., Haug, L. S., Hernandez-Ferrer, C., Heude, B., Karachaliou, M., Krog, N. H., McEachan, R., … Basagaña, X. (2019). Early-life environmental exposures and blood pressure in children: An exposome approach. Journal of the American College of Cardiology, 74(10), 1317–1328. https://doi.org/10.1016/j.jacc.2019.06.069

Wolska, L., Mechlińska, A., Rogowska, J., & Namieśnik, J. (2014). Polychlorinated biphenyls (PCBs) in bottom sediments: Identification of sources. Chemosphere, 111, 151–156. https://doi.org/10.1016/j.chemosphere.2014.03.025

Yu, D., Liu, X., Liu, X., Cao, W., Zhang, X., Tian, H., Wang, J., Xiong, N., Wen, S., Wu, Y., Sun, X., & Zhou, Y. (2019). Polychlorinated Dibenzo-p-Dioxins, Polychlorinated Dibenzofurans, and Dioxin-Like Polychlorinated Biphenyls in Umbilical Cord Serum from Pregnant Women Living Near a Chemical Plant in Tianjin, China. International Journal of Environmental Research and Public Health, 16(12), E2178. https://doi.org/10.3390/ijerph16122178

Zhu, Y., Gomez, J. A., Laufer, B. I., Mordaunt, C. E., Mouat, J. S., Soto, D. C., Dennis, M. Y., Benke, K. S., Bakulski, K. M., Dou, J., Marathe, R., Jianu, J. M., Williams, L. A., Gutierrez Fugón, O. J., Walker, C. K., Ozonoff, S., Daniels, J., Grosvenor, L. P., Volk, H. E., … LaSalle, J. M. (2022). Placental methylome reveals a 22q13.33 brain regulatory gene locus associated with autism. Genome Biology, 23(1), 46. https://doi.org/10.1186/s13059-022-02613-1

