## Supplemental Figures for "Networks of placental DNA methylation correlate with maternal serum PCB concentrations and child neurodevelopment"

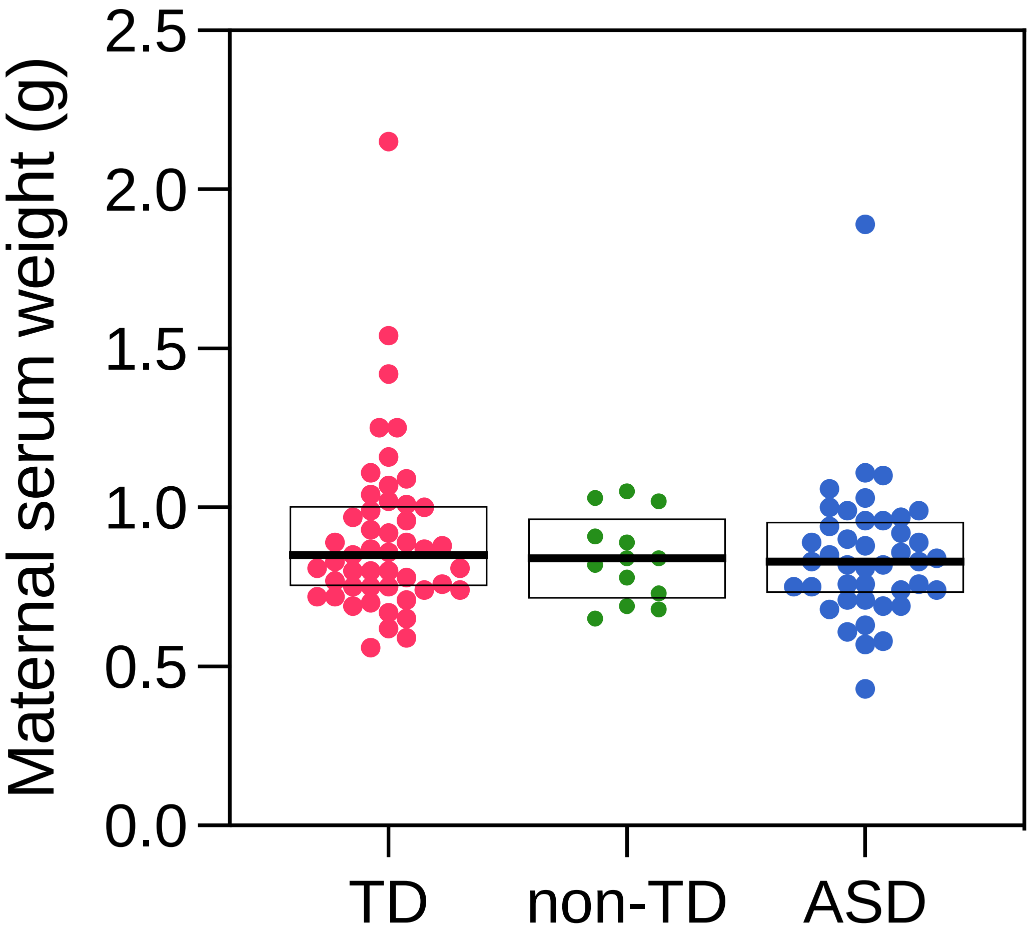


**Figure S1. Maternal serum weight used for PCB measurements does not differ across offspring ASD diagnostic groups.** PCB congeners (ng) were measured in maternal serum samples, which ranged from 0.4259g to 2.1497g. Before adjusting PCB data by serum weight, the serum weights were tested for differences across offspring ASD diagnostic groups (TD, non-TD, ASD) using the Mann-Whitney $U$ test for each pairwise comparison (TD vs non-TD: $U$ = 291, $p$-value = 0.6412; TD vs ASD: $U$ = 927, $p$-value = 0.4196; non-TD vs ASD: $U$ = 269, $p$-value = 0.9415), and the Kruskal-Wallis test for comparison across all three groups (Kruskal-Wallis statistic = 0.7172, $p$-value = 0.6986). No differences were detected, so PCB data were adjusted for serum weight for analysis. Boxes indicate 1$^{st}$ quartile, median, and 3$^{rd}$ quartile.

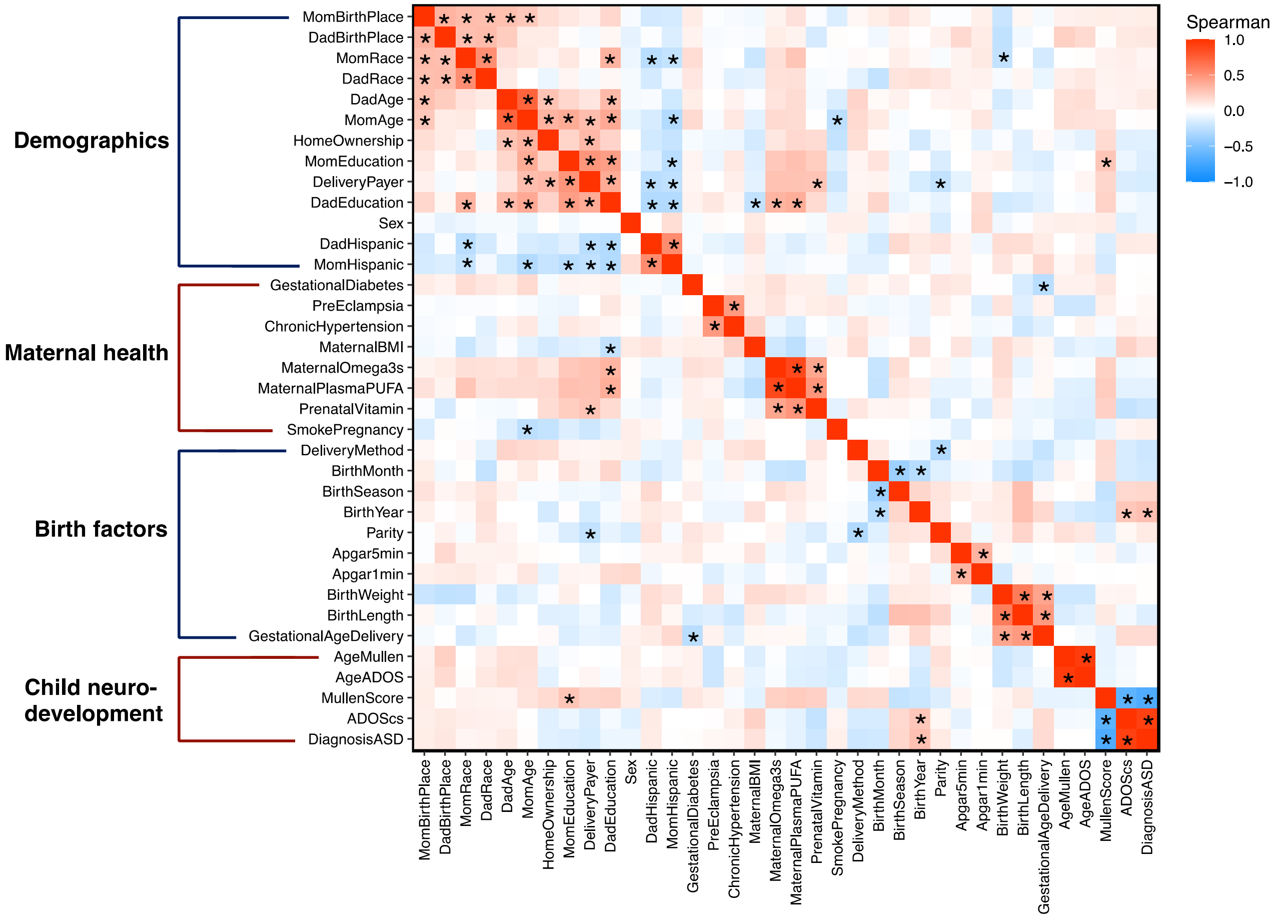


**Fig S2. Trait-trait correlations with Spearman correlation.** Traits were correlated using Spearman correlation to evaluate potential monotonic relationships that were not captured by Pearson correlation (included as a main figure). Traits are grouped into four categories and ordered hierarchically within each category. * FDR < 0.05.

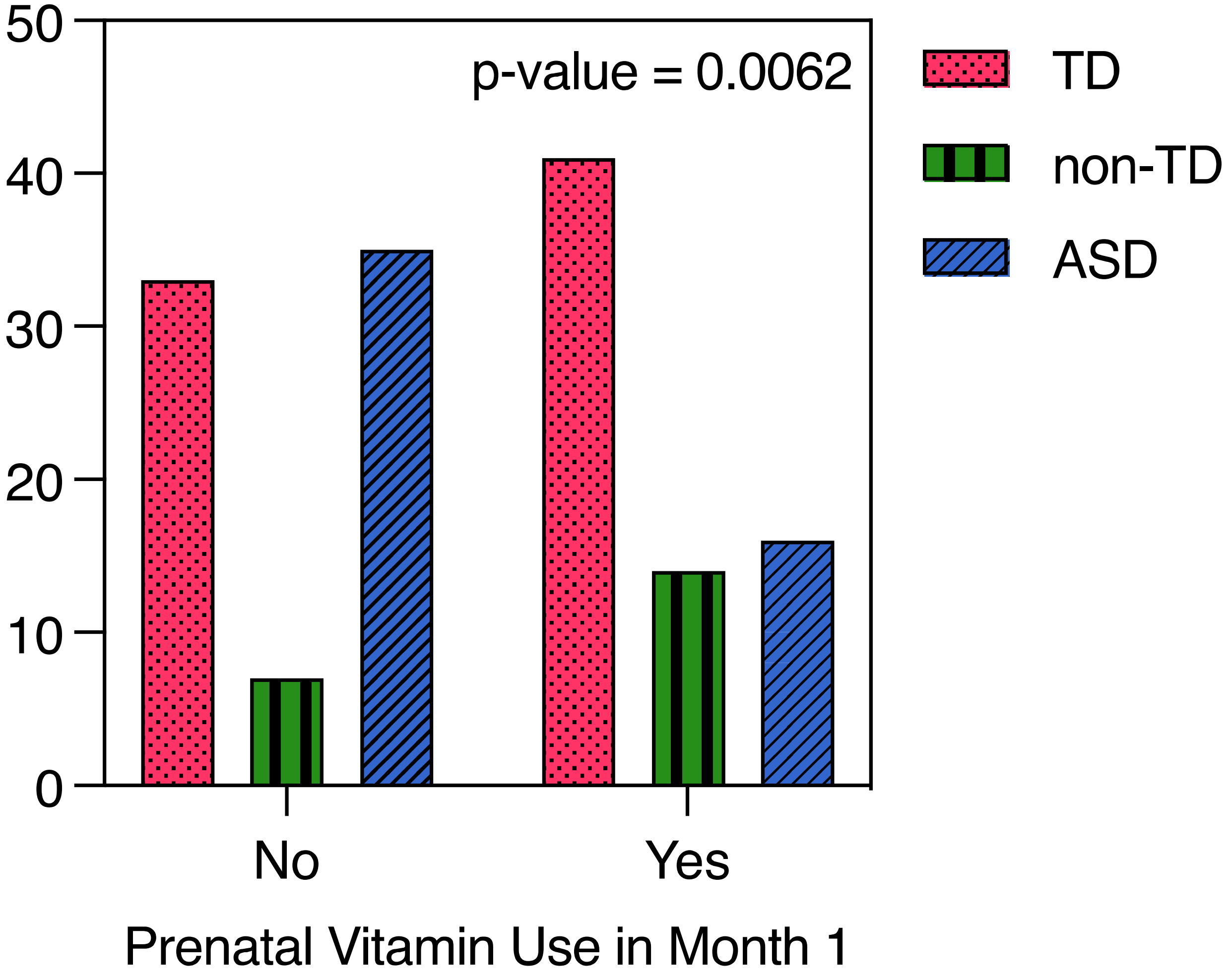


**Fig S3. Diagnosis of ASD in the offspring was less common in the mothers who took prenatal vitamins in month 1 of pregnancy.** In the group of mothers who did take prenatal vitamins in month 1 (*n*=71), there were more TD (*n*=41 (57.7%)) and non-TD (*n*=14 (19.7%)) offspring and fewer ASD (*n*=16 (22.5%)) offspring, compared to the group of mothers who did not take prenatal vitamins (*n*=75, TD *n*= 33 (44.0%), non-TD *n*= 7 (9.3%), ASD *n*= 35 (46.7%)). Bars represent the number of offspring in each diagnostic category separated by prenatal vitamin use in month 1. Chi-square = 10.17, *p*-value = 0.0062.

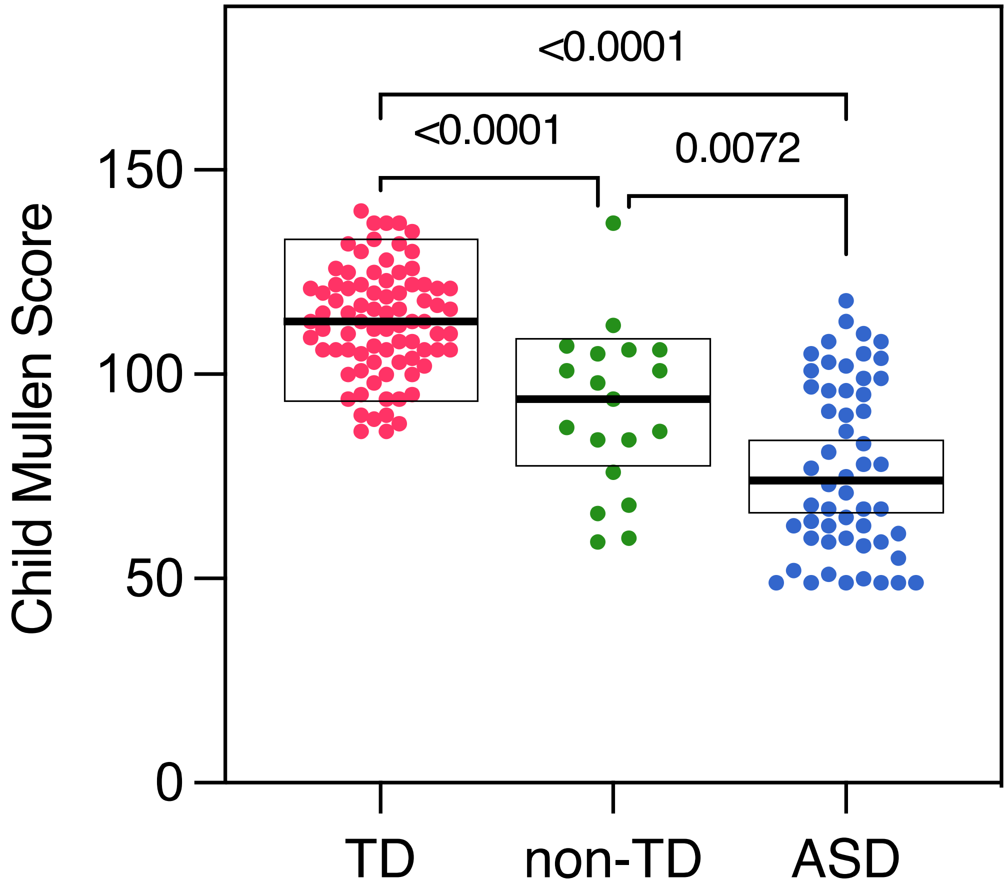


**Fig S4. Child Mullen scores are significantly different across ASD diagnostic groups.** Mullen scores are one component of ASD diagnosis and reflect child cognition. Mullen scores overlapped for each diagnostic group but an analysis of variance (ANOVA) showed significant variation among groups ($p$-value < 0.0001). A post-hoc Tukey multiple comparisons test showed that the means of each group were significantly different (TD vs non-TD $p$-value < 0.0001, TD vs ASD $p$-value < 0.0001, non-TD vs ASD $p$-value = 0.0072). Boxes indicate 1$^{st}$ quartile, median, and 3$^{rd}$ quartile.

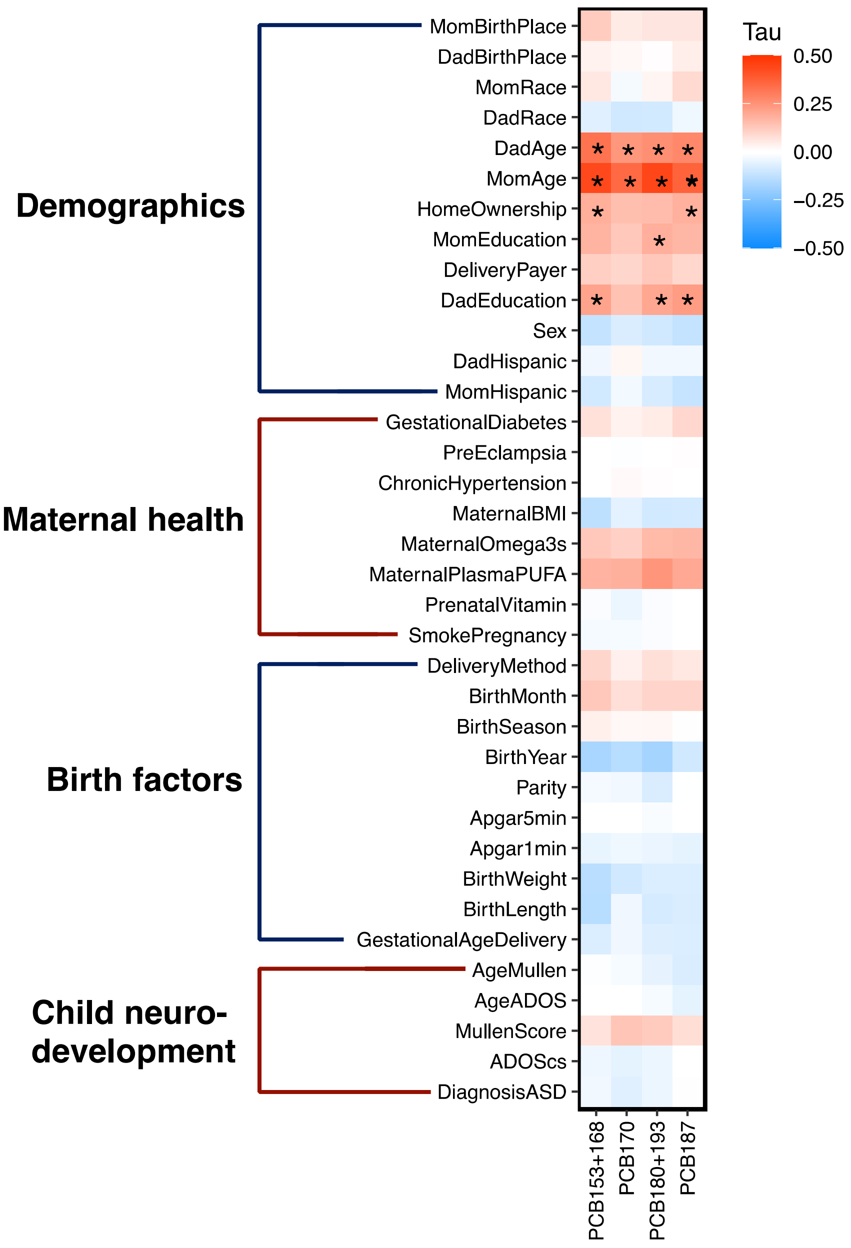


**Fig S5. Sample traits correlated with maternal serum PCB concentrations using Kendall's tau.** Correlations consider censored and uncensored data points from PCB 153 + 168, 170, 180 + 193, 187 (ng p g). Traits are grouped into four categories and ordered hierarchically within each category. The correlation matrix is the same as shown as Figure 3B, except that * indicates FDR < 0.05.

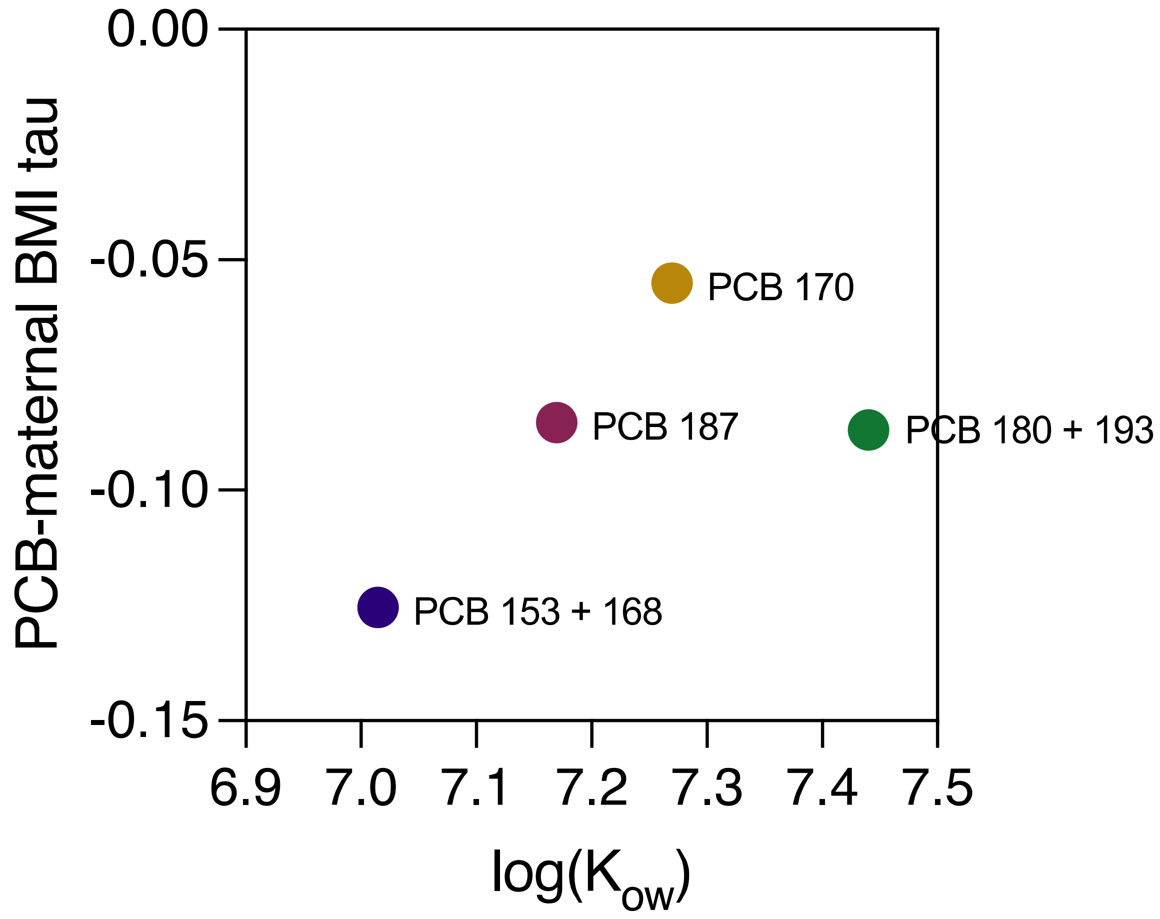


**Fig S6. PCB 153 + 168 is the least lipophilic of the four congeners/congener pairs analyzed, and has the strongest negative PCB-maternal BMI correlation.** The correlation coefficients (Kendall's tau) between maternal BMI and PCB 153 + 168, 170, 180 + 193, and 187 is plotted against log($K_{ow}$), the octanol water coefficients for each PCB congener/congener pair. For congener pairs (153 + 168 and 180 + 193), their $K_{ow}$ values were averaged. Higher log($K_{ow}$) values indicate greater lipophilicity, showing that PCB 153 + 168 is the least lipophilic and would more readily release from fat stores than more lipophilic PCBs. This slightly easier release from fat stores may result in higher concentrations of PCB 153 + 168 in those with low BMI, resulting in a strongly negative PCB-BMI correlation. More lipophilic PCBs may remain in fat stores more readily, even in individuals with low BMI.

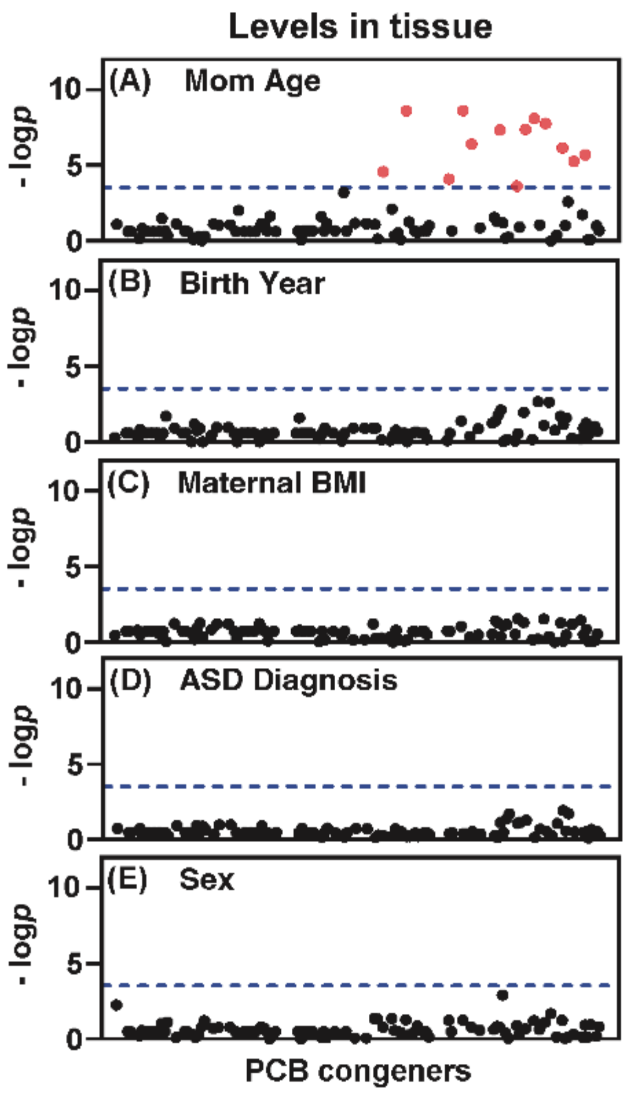


**Fig S7. Levels of single or co-eluting PCB congeners reveal significant differences between mom age groups (A) but not offspring birth year (B), maternal BMI (C), offspring ASD diagnosis (D), or offspring sex (E).** A Tobit model was used to detect differences across groups (mom Age: >35 vs ≤35, offspring birth year: 2006-2011 vs 2012-2016, maternal BMI: >25 vs ≤25, offspring ASD diagnosis: ASD vs non-TD vs TD, offspring sex: male vs female. The dotted line indicates the value of log *p* x (-1) of Bonferroni-adjusted multiple comparisons (*p*-value = 2.89 x 10-4). Red dots indicate PCB congeners with a significant difference. PCB congeners are plotted on the x-axis in the order of their Ballschmiter and Zell number, as defined by the EPA. Congeners with *p*-value = 1 were not plotted. The *p* values of the corresponding PCB congeners are presented in Table S16.

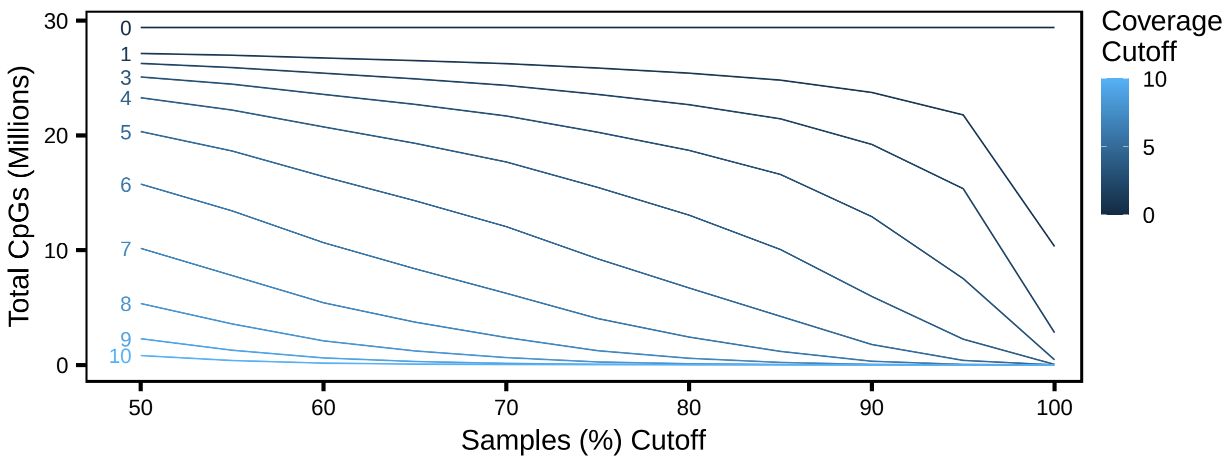


**Fig S8. Comethyl CpG totals are altered by coverage and percent of samples.** Total CpGs decrease as coverage cutoff (number of reads per sample) and percent of samples with that coverage increase. CpG totals are presented in Table S17.

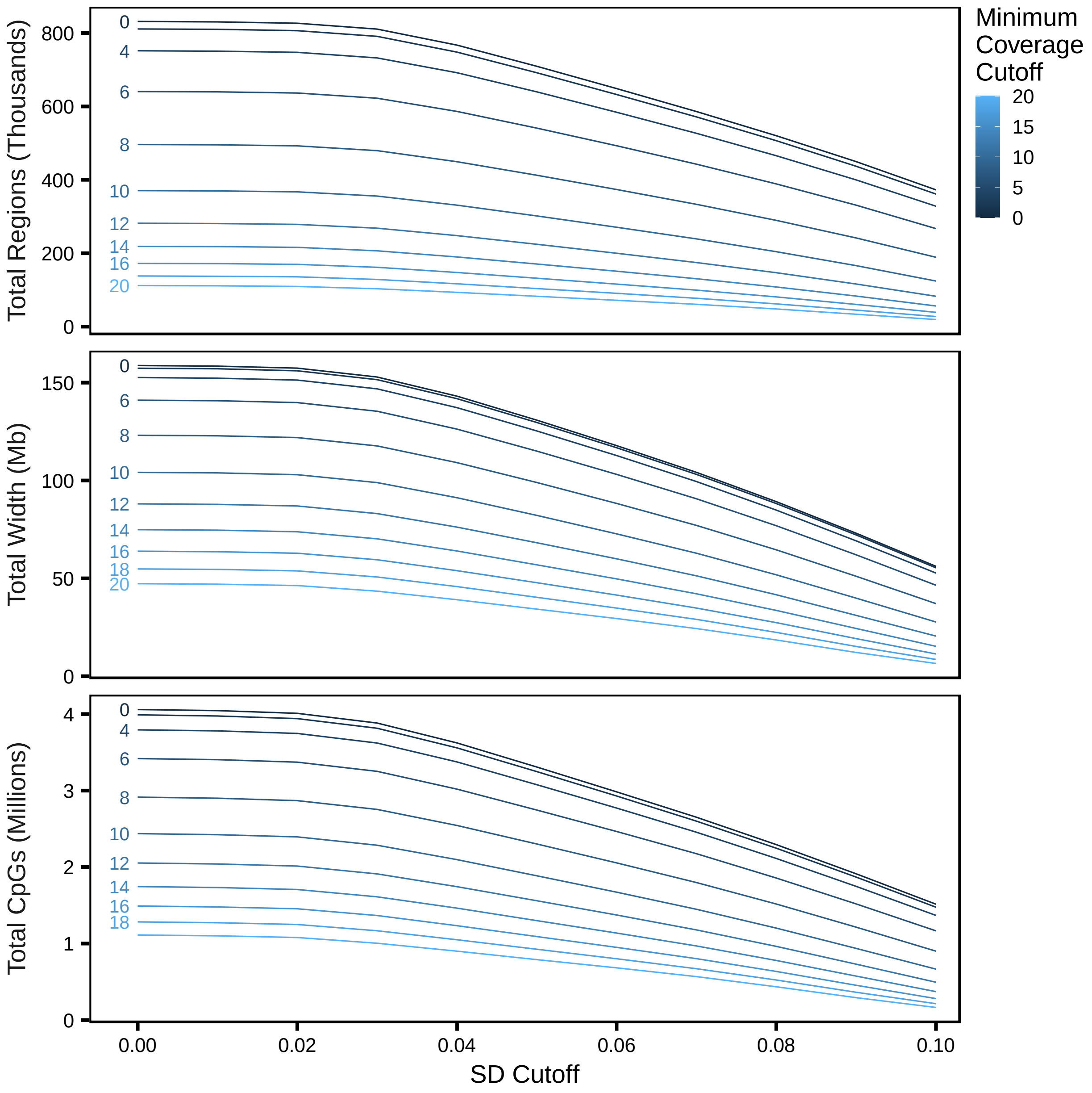


**Fig S9. Comethyl region totals are altered by coverage and standard deviation thresholds.** Number of total regions, total width covered by regions, and number of total CpGs covered by regions, decrease as coverage (minimum reads in all samples) and minimum standard deviation (difference in methylation across samples) increase.

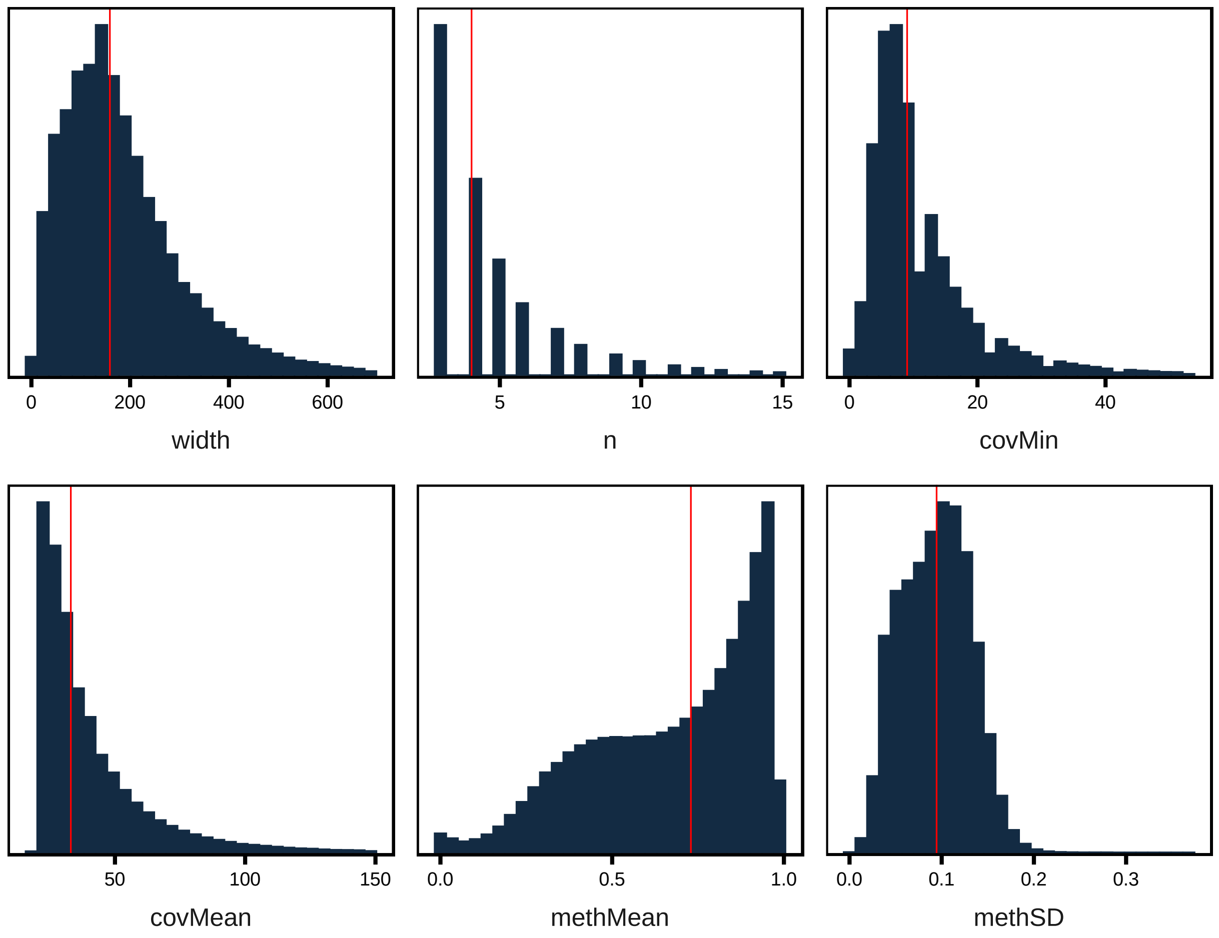


**Fig S10. Comethyl region characteristics before filtering.** Plots show the number of base pairs within each region (width), number of CpGs within each region (n), minimum coverage (covMin), mean coverage (covMean), mean methylation across a region (methMean), and methylation standard deviation across samples for a given region (methSD). The red line indicates the median value.

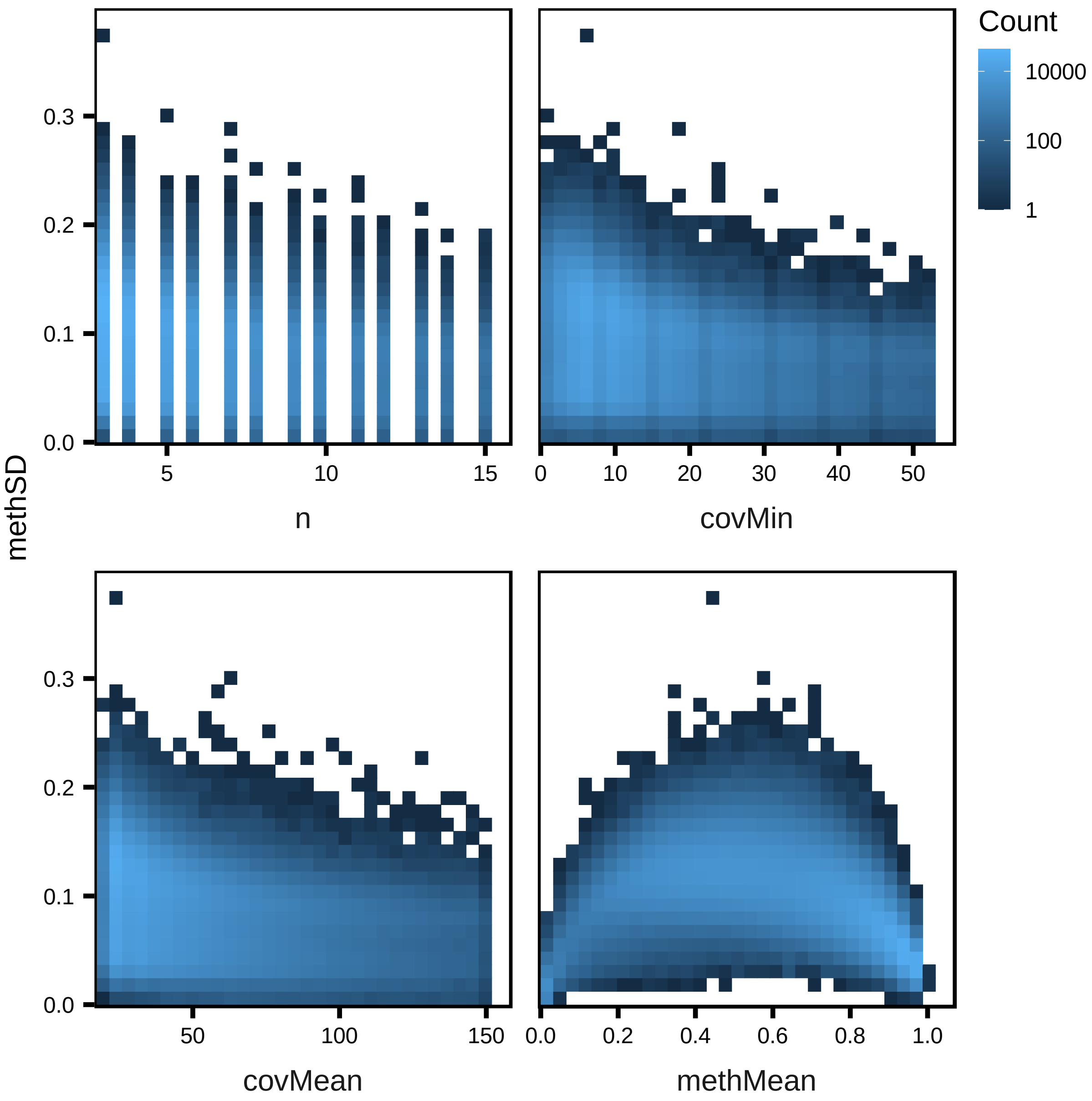


**Fig S11. Comethyl regions characteristics vs methylation standard deviation before filtering.** Methylation standard deviation across samples (methSD) tended to decrease as number of CpGs within each region (n), minimum coverage (covMin), and mean coverage (covMean) increased, but was highest when mean methylation across a region (methMean) was near 0.6.

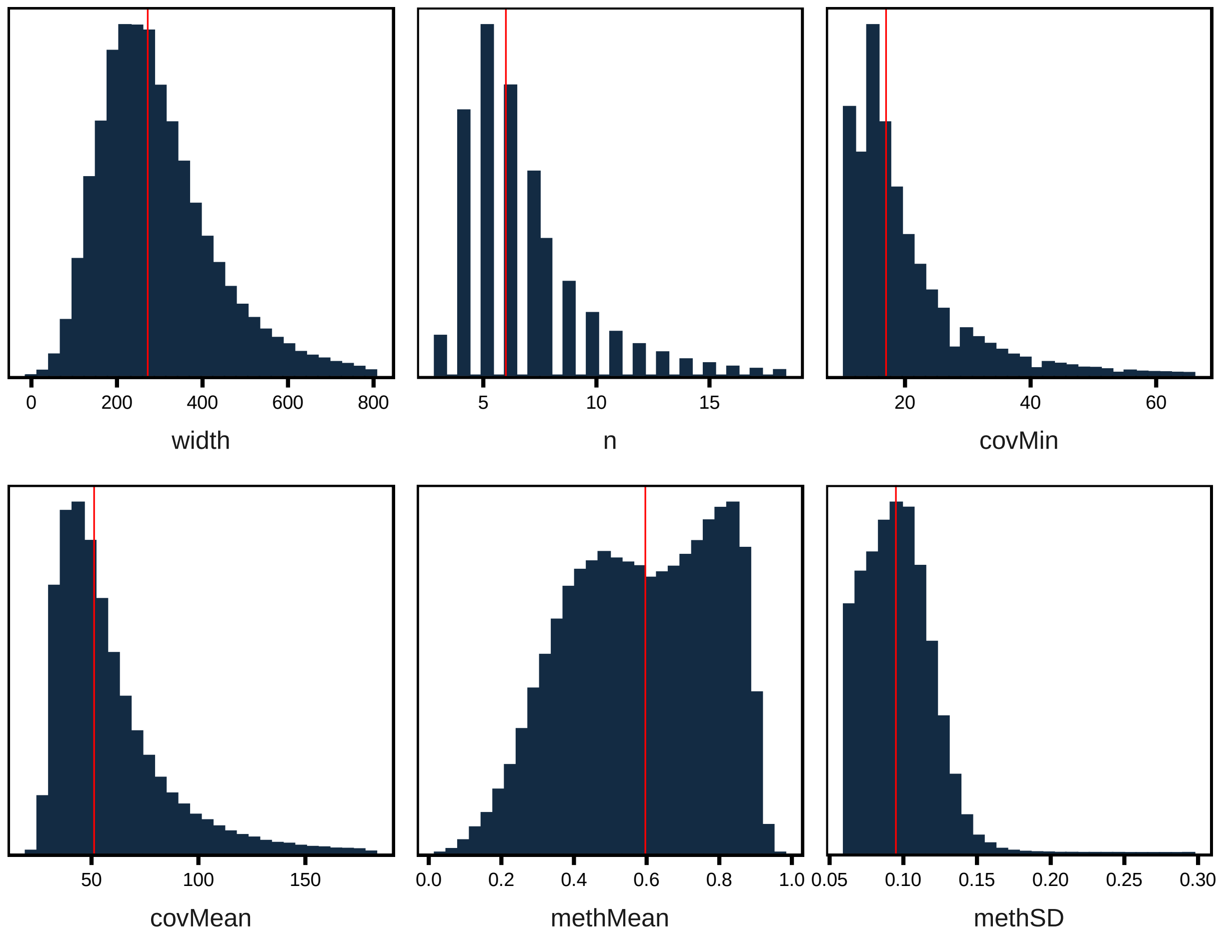


**Fig S12. Comethyl region characteristics after filtering.** Plots show the number of base pairs within each region (width), number of CpGs within each region (n), minimum coverage (covMin), mean coverage (covMean), mean methylation proportion across a region (methMean), and methylation standard deviation across samples for a given region (methSD). The red line indicates the median value. Regions were filtered to those with covMin ≥ 12 and methSD ≥ 0.06. The methMean plot shows the bimodal methylation pattern characteristic of placental tissue, where the hump to the left of the red line indicates a partially methylated domain (PMD), and the hump to the right of the red line indicates a highly methylated domain (HMD).

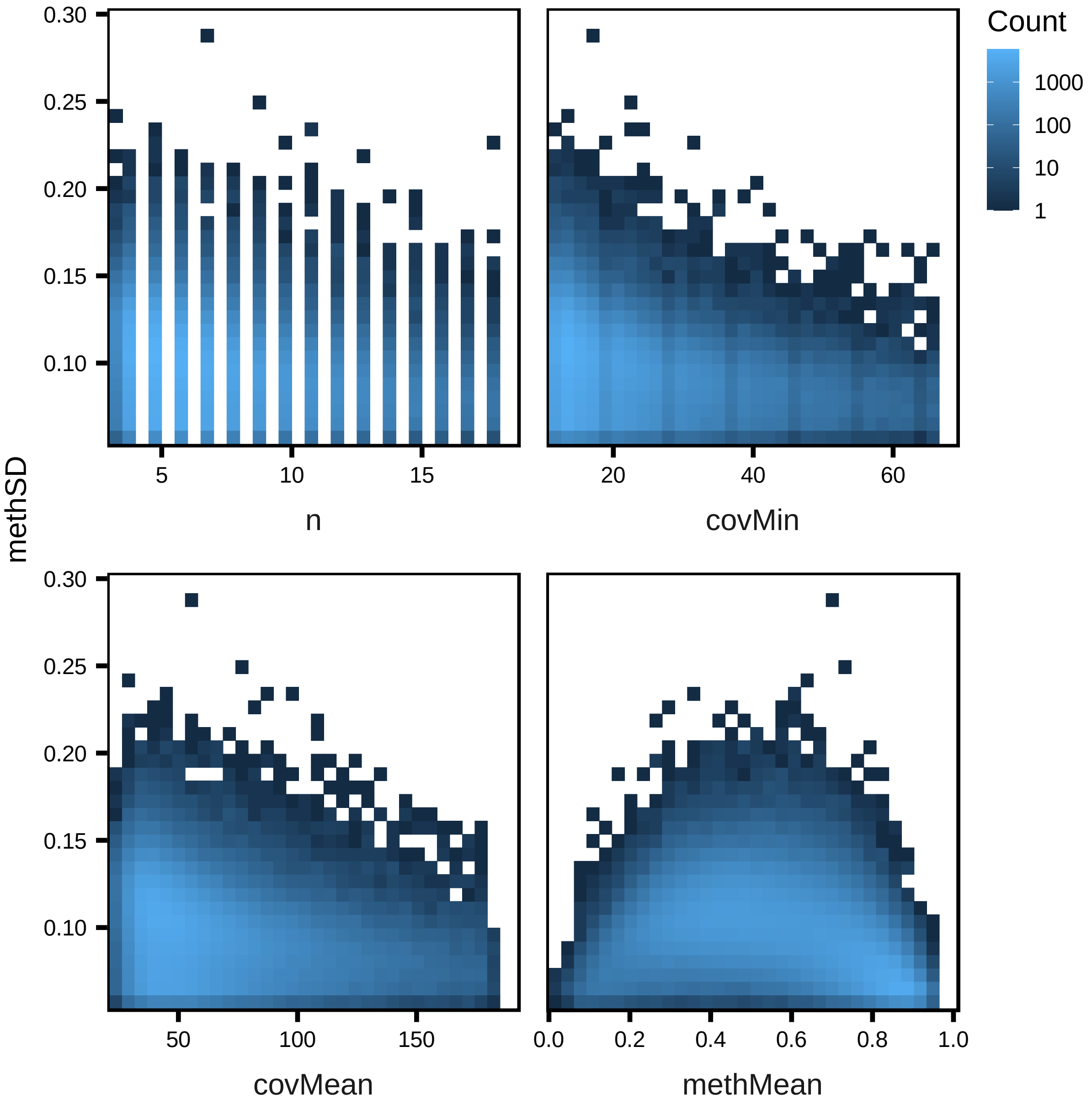


**Fig S13. Comethyl regions characteristics vs methylation standard deviation after filtering.** Methylation standard deviation across samples (methSD) tended to decrease as number of CpGs within each region (n), minimum coverage (covMin), and mean coverage (covMean) increased, but was highest when mean methylation across a region (methMean) was between 0.4 and 0.6, representing partially methylated domains (PMDs).

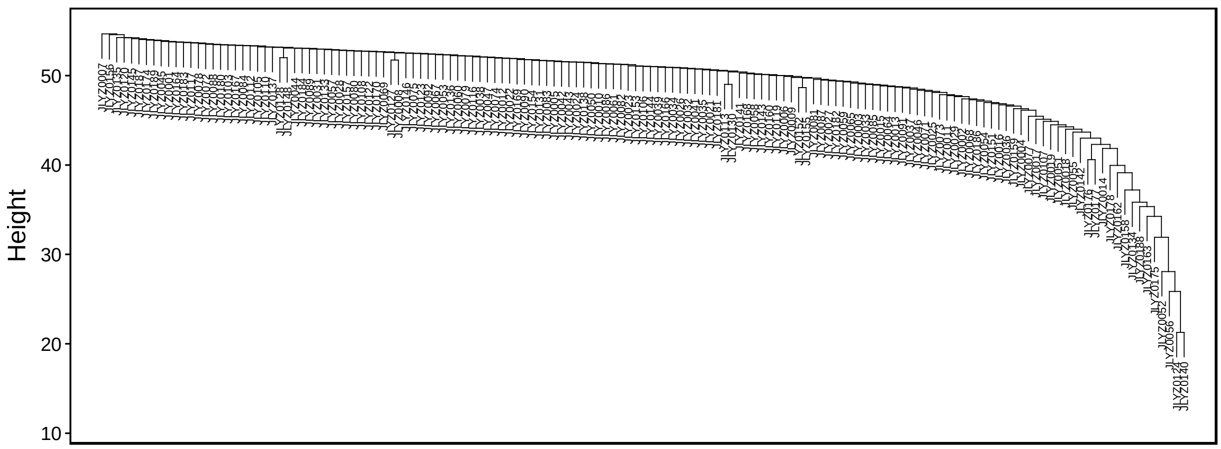


**Fig S14. Comethyl sample dendrogram shows no outliers.** Samples were clustered by Euclidean distance and none were shown to be outliers.

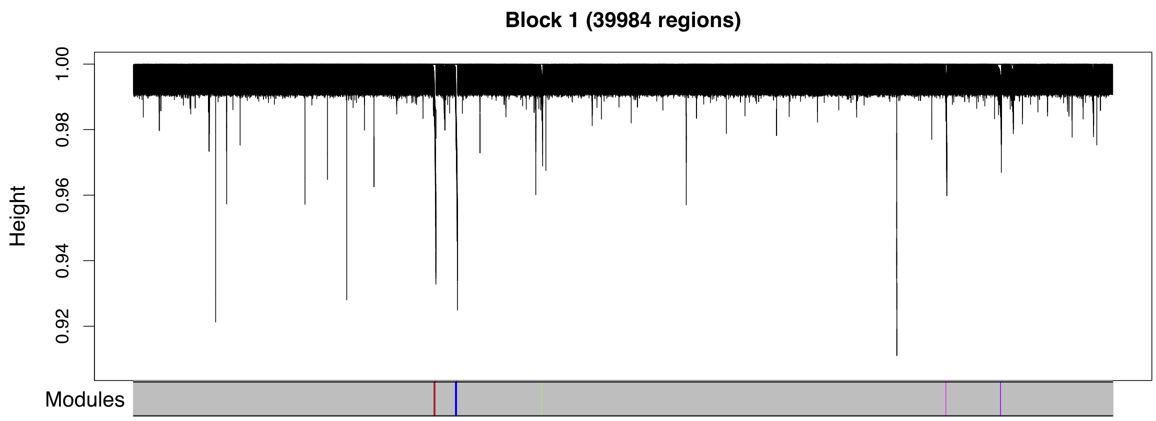

Block 1 (39984 regions)
Height
1.00
0.98
0.96
0.94
0.92
Modules


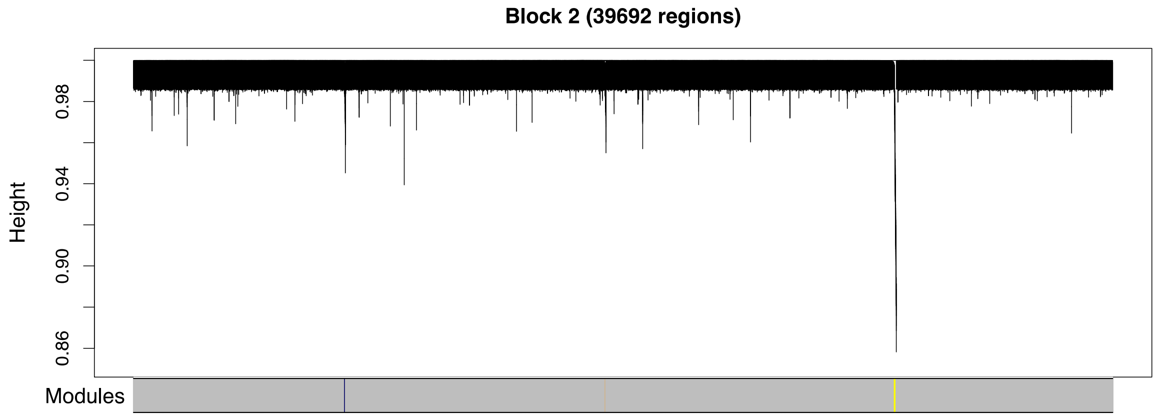

Block 2 (39692 regions)
Height
0.98
0.94
0.90
0.86
Modules


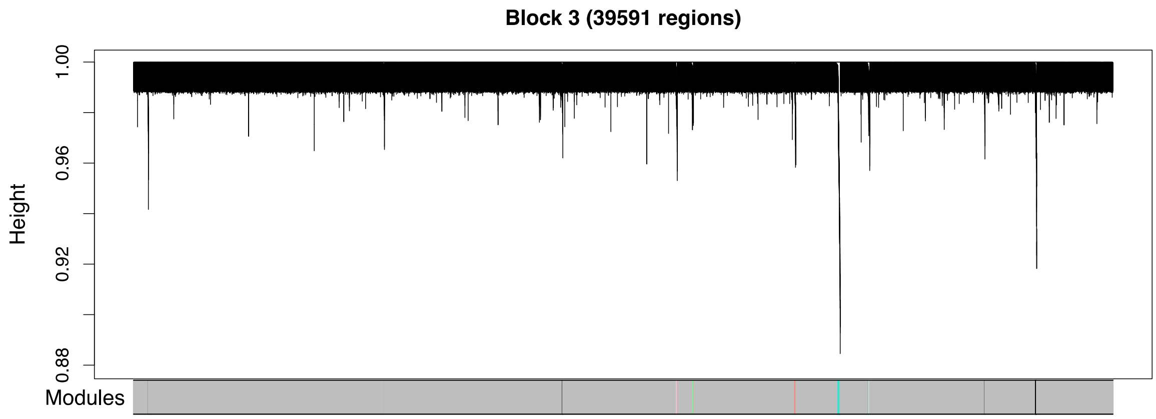

Block 3 (39591 regions)
Height
1.00
0.96
0.92
0.88
Modules

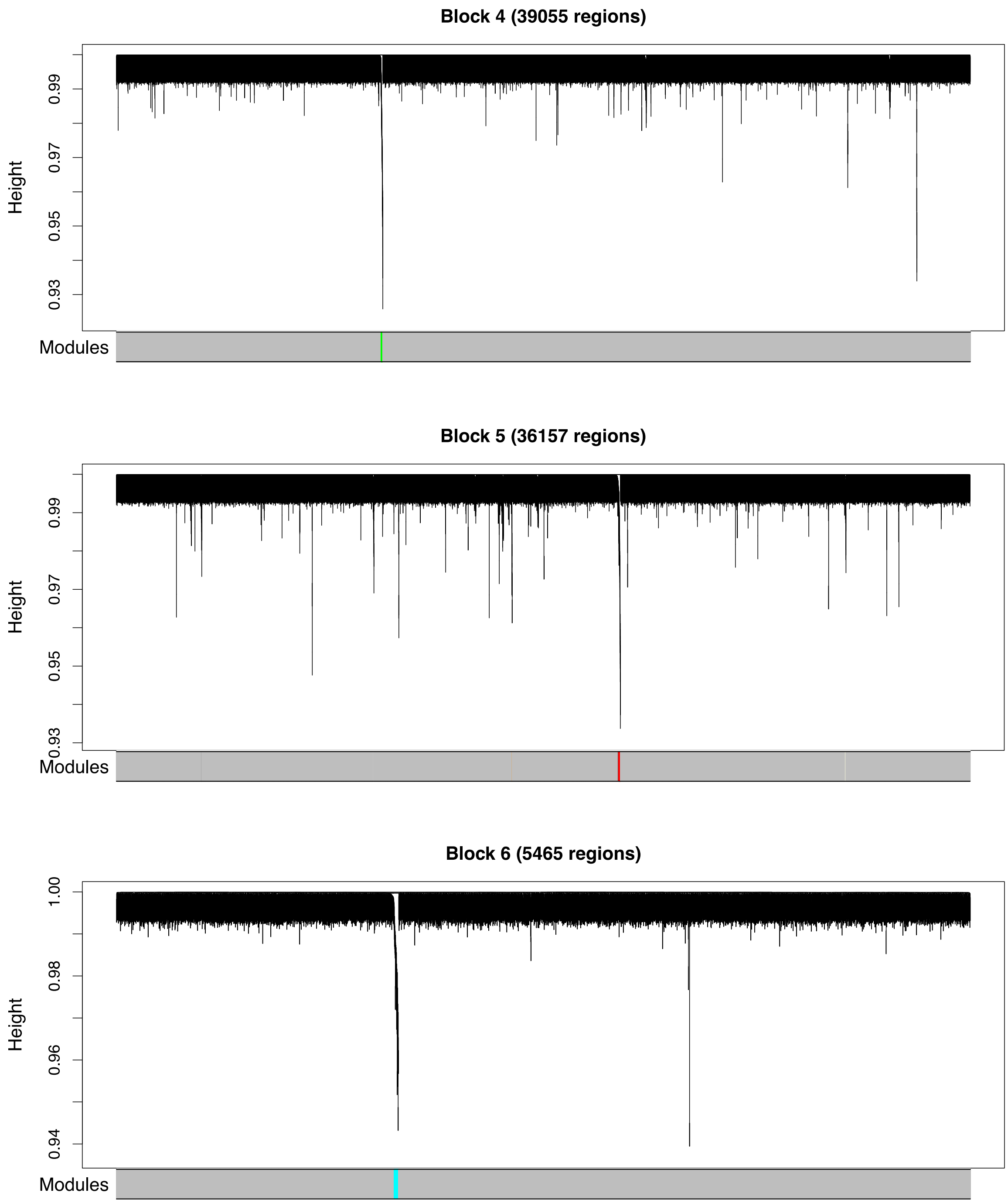


**Fig S15. Comethyl region dendrograms for the six blocks used for network construction.** Regions were formed into blocks close to but not exceeding 40,000 regions. Each block underwent a full network analysis and regions were assigned to modules. If modules from different blocks had highly correlated eigennodes (r>0.9), they were merged into a single module. Regions are clustered based on dissimilarity (1-correlation). Modules were limited to 10 or more regions, where each region consists of 3 or more CpGs separated by no more than 150 bp.

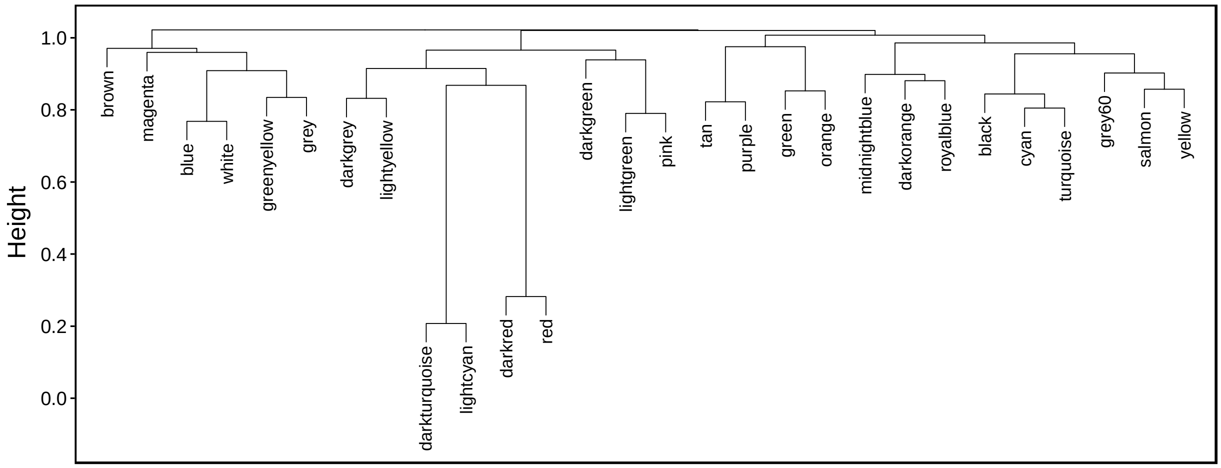


**Fig S16. Comethyl module dendrogram shows modules clustered by dissimilarity.** Similar modules have lower dissimilarity (height), such as darkturquoise with lightcyan and darkred with red.

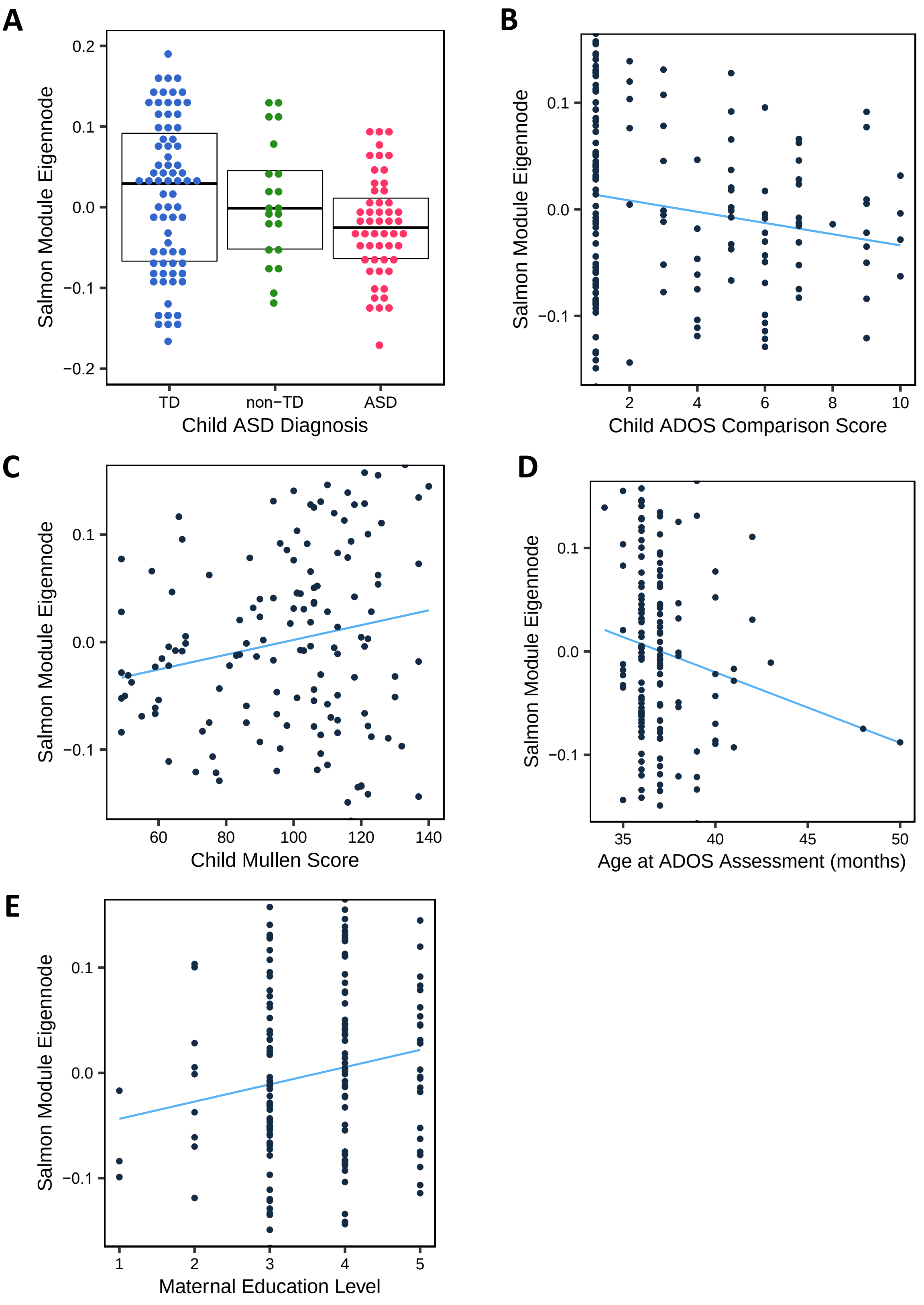


**Fig S17. *GALC* Salmon module eigennodes significantly correlated** with **A)** child ASD diagnosis ($P < 0.010$), **B)** child ADOS comparison score ($P = 0.035$), **C)** child Mullen Score ($P = 0.037$), **D)** child age at ADOS assessment in months ($P = 0.044$), and **E)** maternal education level, where 1 = less than high school, 2 = high school diploma / GED, 3 = some college, 4 = Bachelor's degree, and 5 = graduate or professional degree ($P = 0.029$)

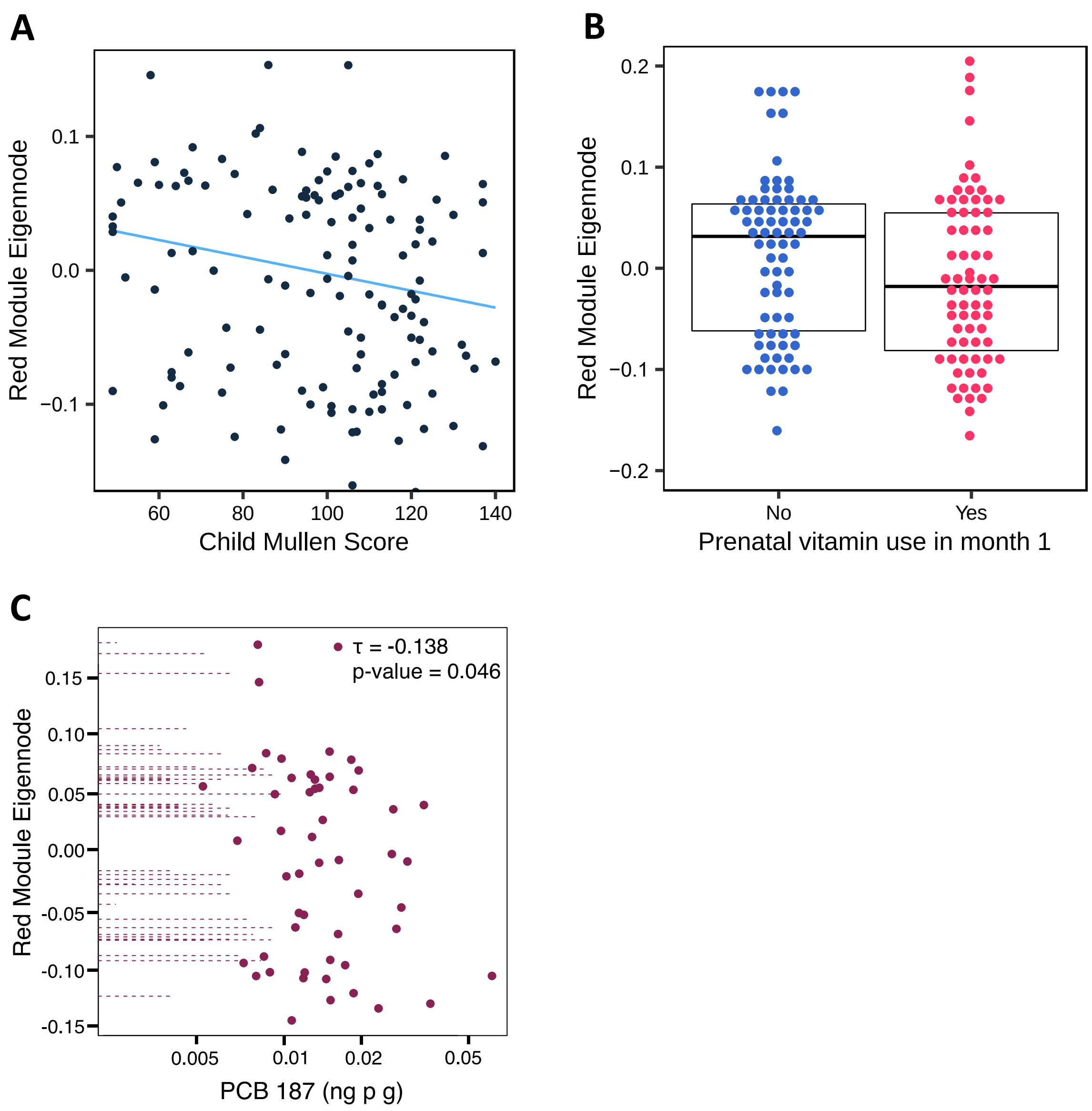


**Fig S18.** ***AUTS2*** **Red module eigennodes significantly correlated** with **A)** child Mullen Score ($P$ = 0.040) **B)** maternal prenatal vitamin use in month 1 of pregnancy ($P$ = 0.048) and **C)** maternal serum levels of PCB 187 (ng p g) ($P$ = 0.046)

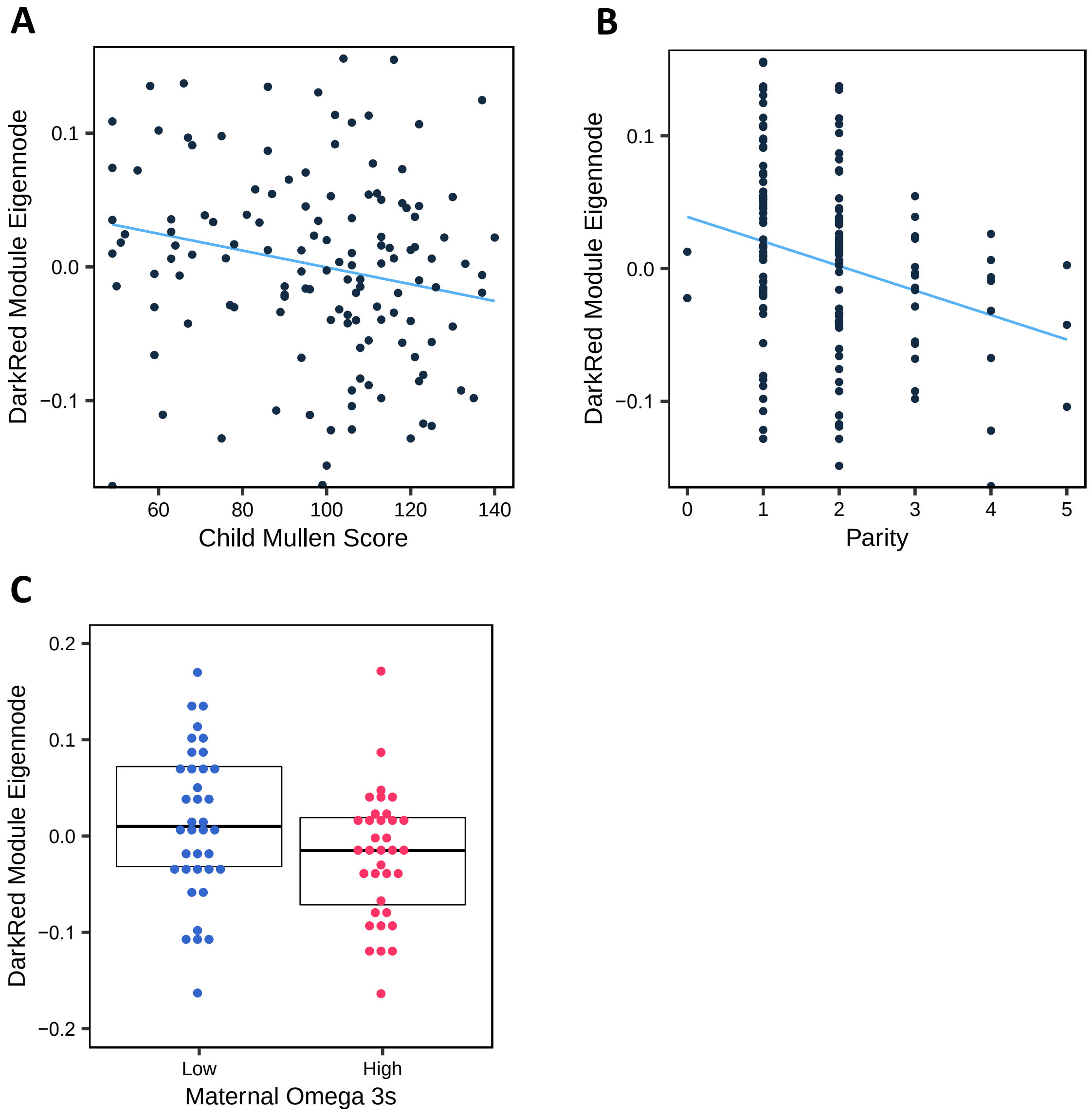


**Fig S19.** ***AUTS2*** **Darkred module eigennodes significantly correlated** with **A)** child Mullen Score (*P* = 0.046) **B)** parity (number of pregnancies the mother has had over 20 weeks gestation prior to the child in this study) (*P* = 0.003) **C)** maternal omega 3 levels from third trimester plasma samples (*P* = 0.036)

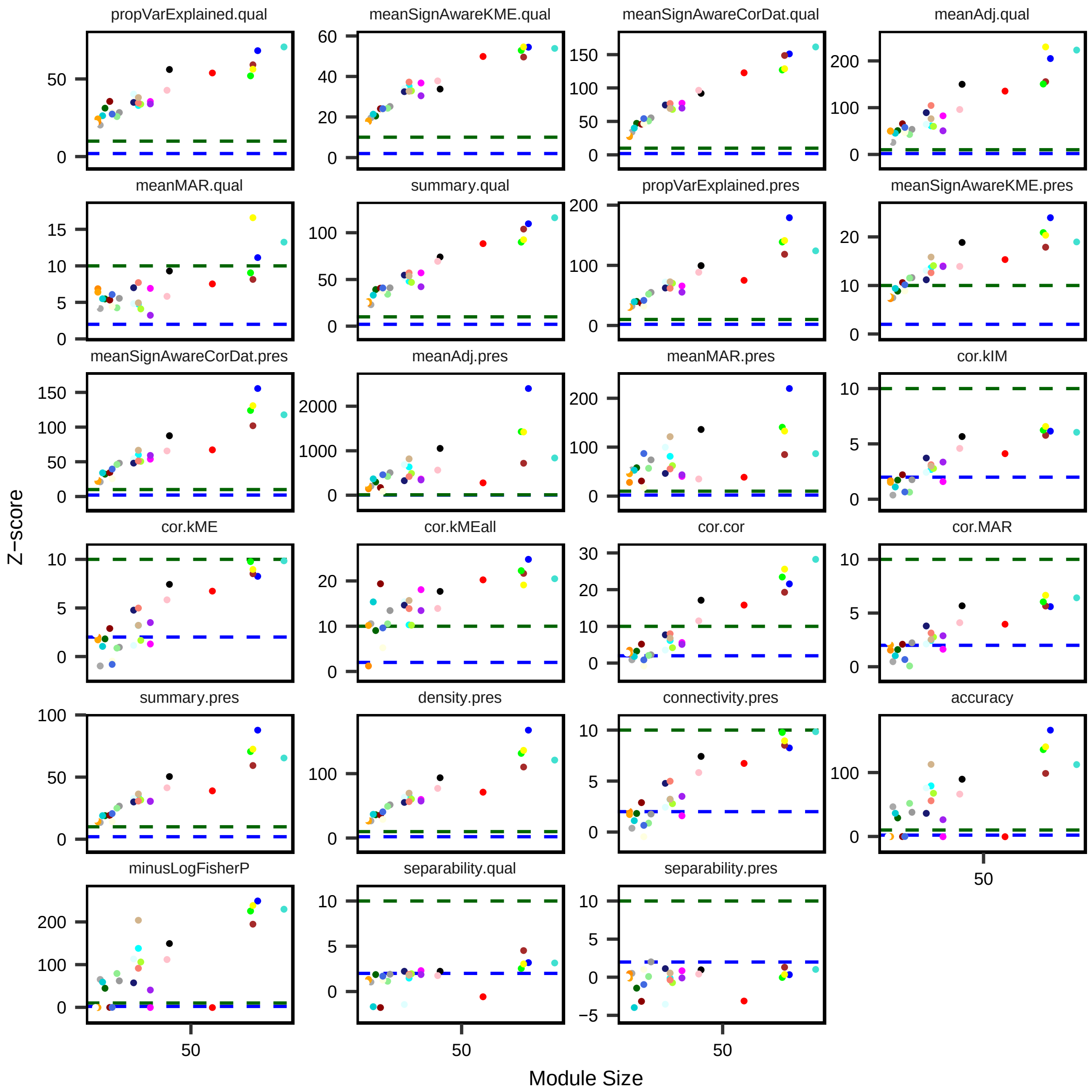


**Fig S20. Comethyl module quality and preservation statistics.** An independent dataset of placenta samples was used to build a comethylation network using the same set of CpGs and regions. Statistics related to quality and preservation were calculated based on the two networks and compared to an empirical null distribution to generate Z-scores. Blue and green dotted lines represent weak/moderate and strong evidence, respectively.
